# Genetic Architecture and Myocardial Fibrotic Remodeling in Mitral Valve Prolapse

**DOI:** 10.64898/2026.04.09.26350328

**Authors:** Aeron M Small, Mengyao Yu, Takiy E Berrandou, Adrien Georges, Matthew Huff, Jordan E Morningstar, Søren A Rand, Satoshi Koyama, Jiwoo Lee, Ha My Vy, Eric Farber-Eger, Shilin Jin, Maja-Theresa Dieterlen, Amy R Kontorovich, Ta-Yu Yang, Ron Do, Martina Dreßen, Markus Krane, Nina Feirer, Stefanie A Doppler, Heribert Schunkert, Teresa Trenkwalder, Quinn S Wells, Klaus Berger, Sisse R Ostrowski, Erik Sørensen, Ole B Pedersen, Johan S Bundgaard, Jonas Ghouse, Henning Bundgaard, Andrea Ganna, Christian Erikstrup, Christina Mikkelsen, Mie T Bruun, Bitten Aagaard, Henrik Ullum, Erik Abner, Susan A Slaugenhaupt, Lincoln Nadauld, Kirk Knowlton, Anna Helgadottir, Gardar Sveinbjornsson, Daniel F Gudbjartsson, Pall Melsted, Francesca Delling, Michael Rosenberg, Lohit Garg, Christopher Gignoux, Renae Judy, Michael G Levin, Scott M Damrauer, Patrick Ellinor, David Milan, Robert A Levine, Michael Borger, James C Engert, George Thanassoulis, Peter WF Wilson, Kelly Cho, VA Million Veteran Program, Gina M Peloso, Russell A Norris, Nabila Bouatia-Naji, Pradeep Natarajan

**Affiliations:** Division of Cardiology, Department of Medicine, VA Boston Healthcare System, Boston, MA, USA; Heart and Vascular Institute, Mass General Brigham, Boston, MA, USA; Program in Medical and Population Genetics and the Cardiovascular Disease Initiative, Broad Institute of Harvard and MIT, Cambridge, MA, USA; Human Phenome Institute, Shanghai Pudong Hospital, Fudan University, Shanghai, China; Univ Paris Cite, Inserm, PARCC, Paris, France; Department of Regenerative Medicine and Cell Biology, Medical University of South Carolina, Charleston, SC, USA; Department of Medicine, Medical University of South Carolina, Charleston, SC, USA; Department of Cardiology, Rigshospitalet, Copenhagen University Hospital, Copenhagen, Denmark; Broad Institute of MIT and Harvard, Cambridge, MA, USA; Instiute for Molecular Medicine Finland, HiLIFE, University of Helsinki, Helsinki, Finland; Department of Genetics and Genomic Sciences, Icahn School of Medicine at Mt. Sinai, New York City, New York, USA; Division of Cardiovascular Medicine, Vanderbilt University Medical Center, Nashville, TN, USA; Shanghai Pudong Hospital, Fudan University, Shanghai, China; Department of Cardiac Surgery, HELIOS Clinic, Heart Center Leipzig, University Hospital Leipzig, Leipzig, Germany; Mount Sinai Fuster Heart Hospital, New York City, New York, USA; McGill University, Montréal, Quebec, Canada; Department of Artificial Intelligence and Human Health, Icahn School of Medicine at Mt. Sinai, New York City, New York, USA; The Charles Bronfman Institute for Personalized Medicine, Icahn School of Medicine at Mt. Sinai, New York City, New York, USA; Department of Cardiovascular Surgery, Institute Insure, TUM University Hospital German Heart Center, TUM School of Medicine & Health, Technical University of Munich, Munich, Germany; German Center for Cardiovascular Research (DZHK), Partner site Munich Heart Alliance, Munih, Germany; Department of Cardiology, TUM University Hospital German Heart Center,, TUM School of Medicine & Health, Technical University of Munich, Munich, Germany; Institute of Epidemiology and Social Medicine, University of Münster, Munster, Germany; Department of Clinical Immunology, Copenhagen University Hospital, Rigshospitalet, Copenhagen, Denmark; Department of Clinical Medicine, Faculty of Health and Medical Sciences, University of Copenhagen, Copenhagen, Denmark; Department of Cardiology, Copenhagen University Hospital, Rigshospitalet, Copenhagen, Denmark; Department of Clinical Immunology, Aarhus University Hospital, Aarhus, Denmark; Analytical and Translational Genetics Unit, Massachusetts General Hospital, Harvard Medical School, Boston, MA, USA; Department of Clinical Medicine, Aarhus University Hospital, Aarhus, Denmark; Clinical Immunology Research Unit, Department of Clinical Immunology, Odense University Hospital, Odense, Denmark; Department of Clinical Research, University of Southern Denmark, Odense, Denmark; Department of Clinical Immunology, Aalborg University Hospital, Aalborg, Denmark; Statens Serum Institut, Copenhagen, Denmark; Estonian Genome Center, Institute of Genomics, University of Tartu, Tartu, Estonia; Center for Genomic Medicine, Massachusetts General Hospital and Department of Neurology, Harvard Medical School, Boston, MA, USA; Intermountain Healthcare, St. George, UT, USA; Intermountain Healthcare, Salt Lake City, UT, USA; Amgen deCODE genetics, Inc., Sturlugata 8, Reykjavik 101, Iceland; Department of Medicine, Cardiovascular Division, University of California, San Francisco, CA, USA; Colorado Center for Personalized Medicine, University of Colorado Anschutz Medical Campus, Aurora, CO, USA; Division of Cardiovascular Medicine, Department of Medicine, University of Pennsylvania Perelman School of Medicine, Philadelphia, PA, USA; Corporal Michael Crescenz VA Medical Center, Philadelphia, PA, USA; Department of Surgery, Perelman School of Medicine, University of Pennsylvania, Philadelphia, PA, USA; Leducq Foundation, Boston, MA, USA; University Clinic of Cardiac Surgery, Leipzig Heart Center, Leipzig, Germany; McGill University Health Centre and Research Institute, Montréal, Quebec, Canada; Atlanta VA Medical Center, Atlanta, GA, USA; Department of Medicine, Harvard Medical School, Boston, MA, USA; Million Veteran Program (MVP) Coordinating Center, Veterans Affairs Healthcare System, Boston, MA; Department of Medicine, Division of Aging, Brigham and Women’s Hospital, Boston, MA, USA; Department of Biostatistics, Boston University School of Public Health, Boston, MA, USA; Million Veteran Program (MVP) Coordinating Center, Veterans Affairs Healthcare System, Boston, MA, USA; Center for Genomic Medicine, Massachusetts General Hospital, Boston, MA, USA

**Author notes:** These authors contributed equally (Aeron M Small, Mengyao Yu, Takiy E Berrandou, Adrien Georges). These authors contributed equally (Nabila Bouatia-Naji, Pradeep Natarajan).

## Abstract

Mitral valve prolapse (MVP) is the most common cause of primary mitral regurgitation and is associated with the development of malignant arrhythmias, often in the context of myocardial fibrosis. The genetic architecture of MVP, and whether there are genetic factors explaining why only some individuals with MVP have adverse outcomes, remains poorly understood. We performed a meta-analysis of genome-wide association studies (GWAS) for MVP encompassing 21,517 cases among a total sample size of over 2.2 million individuals. We discovered 89 genomic risk loci for MVP, of which 72 were novel findings. Prioritization of causal genes and pathways using epigenetic and transcriptomic data from mitral valve and extra-valvular tissues replicated known gene associations to MVP including those involved in TGF-β signaling and extracellular matrix biology, but additionally emphasized a role in MVP for biological pathways relevant to cardiomyocyte biology. Accordingly, we identified several MVP risk loci with pleiotropy to cardiomyopathies, especially hypertrophic cardiomyopathy, and demonstrated a significant genetic correlation between MVP and hypertrophic cardiomyopathy. Finally, we interrogated snRNA-seq data in human papillary muscle tissue from two individuals with severe MVP, characterizing genes associated with both risk of papillary muscle fibrosis and MVP.

## Introduction

Mitral valve prolapse (MVP) is the most common primary mitral valve disorder, characterized by myxomatous degeneration or fibroelastic deficiency of the valve leaflets resulting in abnormal bulging of leaflets into the left atrium during systole^1^. MVP may also be accompanied by elongation of the chordae tendinae and/or papillary muscle fibrosis^1^. MVP affects up to 3% of the general population^2^. While most individuals with MVP have a normal life expectancy, up to 20% develop MVP-related complications^3^, ranging from benign symptoms (i.e., chest discomfort, palpitations) to acute severe mitral regurgitation, infective endocarditis, heart failure, significant arrhythmias, or sudden cardiac death (SCD)^4^. The etiologic basis for MVP is incomplete described. However, even when recognized early, no preventive therapies exist to forestall severe complications. Further, individuals with severe MVP-related complications may ultimately require mitral valve replacement or repair, which entails substantial cost and attendant morbidity and mortality^5^.

There are several established genetic associations in MVP. Individuals with certain Mendelian connective tissue disorders, such as Marfan or Loeys-Dietz syndromes^1^ show enrichment of MVP, and some families have an autosomal dominant transmission of MVP to offspring^6^. However, established monogenic conditions do not appear to comprise the majority of MVP cases. Genome-wide association studies (GWAS) along with familial studies^7-9^ have yielded insights into the genetic basis of MVP. A prior GWAS of approximately 5,000 non-syndromic MVP cases identified 16 significant loci^10,11^, implicating transforming growth factor beta (TGF-β) signaling as well as genes involved in cytoskeletal function and cardiomyopathy as genetic risk factors for MVP. These findings support results from familial and targeted sequencing studies, which prioritize additional genes relevant to TGF-β signaling (e.g., *LTBP2*^*9*^) and cardiomyopathy (e.g. *MYH6^12^*). MVP may co-occur with pathologic myocardial fibrosis, which is linked to malignant arrhythmias and SCD^13^. It is unknown whether there are shared genetic factors between MVP and myocardial fibrosis beyond TGF-β signaling that may predispose individuals to future life-threatening arrhythmic events.

In the present study, we perform a large GWAS meta-analysis of MVP and integrate the results with epigenetic and single-nucleus RNA sequencing (snRNA-seq) data from the human mitral apparatus to comprehensively characterize causal genes and pathways. We compare cell-specific gene expression between fibrosed and normal myocardial tissue from individuals with MVP to identify shared biologic pathways between MVP and myocardial fibrosis.

## Methods

### Study populations and phenotyping

The Mitral Valve Prolapse Genetics Consortium (MVP-Gen) comprises 10 studies. Descriptive characteristics for contributing studies are detailed in **Supplemental Table 1** and **Supplemental Methods: Study Descriptions**. MVP was defined using a combination of *International Classification of Diseases* (ICD-10) codes and, where available, echocardiographic diagnosis or documentation of MVP in clinical notes (**Supplemental Table 2**). A detailed description of quality control, imputation, and GWAS methods is provided by study in the **Supplemental Methods**. All participants previously provided informed consent for study procedures with secondary data analysis approved by the Institutional Review Board protocols at sites of analysis.

### Meta-analysis

GWAS summary data from the 10 MVP-Gen contributing sites were uploaded to a central server at the Broad Institute (Cambridge, MA). MVP-Gen studies were meta-analyzed with publicly available MVP GWAS summary data from the recently published MVP GWAS by Roselli et al^10^, which consists of 6 additional, non-overlapping studies. Centrally, study-specific quality control excluded variants with imputation quality ≤ 0.3 and/or a minor allele count of < 10 prior to meta-analysis. Summary statistics in hg19 were converted to hg38 using liftOver v1.04.00^14^. Linkage disequilibrium (LD) score regression intercepts were calculated for all study-specific GWAS summary statistics using LD Score v.1.0.1^15^ and corrected standard errors (SEldsc) were calculated by multiplying the standard error by the square root of the LD score regression intercept when the LD score regression intercept was ≥ 1. Fixed effects, inverse-variance weighted meta-analysis was performed using GWAMA v2.2.2^16^ with SEldsc to correct for inflation. GWAS meta-analysis was performed for the entire multi-ancestry population as well as for ancestry-stratified populations. Variants present in only one study or with a minor allele frequency (MAF) of ≤ 1% were removed from the resulting meta-analysis summary files. Genome-wide significance (GWS) was defined as a P-value < 5 × 10^-8^. Independent lead variants in each GWAS were established by determining the top-most significant variant within a ± 500kb region of each lead variant.

We additionally performed conditional and joint analysis (COJO) using GCTA v1.94.1^17^ to identify secondary independent signals and assess secondary associations within loci of interest. Independent signals were selected using the stepwise model selection procedure (--cojo-slct) with a 2 Mb window around each lead SNP. Genotype data from 10,000 unrelated individuals of European genetic ancestry from the UK Biobank (UKB) were used as the LD reference panel, applying a MAF threshold of 0.01. Conditional analyses were performed on each identified independent SNP to test for additional secondary signals, with genome-wide significance defined as P_Cond_ ≤ 5 × 10^−8^. To ensure that independent secondary signals were robust across genetic ancestries, we additionally evaluated for linkage disequilibrium using the LDpair tool (https://ldlink.nih.gov/?tab=ldpair) between lead variants and secondary independent signals, excluding secondary signals if r^2^ > 0.1 in either African, European, or admixed American reference populations.

### Identification of candidate variants

To generate a list of candidate functional variants for causal variant and gene prioritization, we identified a 95% credible set of variants using the ppfunc function of the corrcoverage package v1.2.1^18^ in R. A posterior probability of causality was evaluated from marginal Z-scores for all variants within 500kb of the lead SNP at each locus. Variants with a cumulated posterior probability of up to 95% were kept for downstream analysis. To consider potentially poorly imputed variants in any individual case-control study, we also included variants in high LD (r^2^>0.8) with the lead SNP at each locus based on a European reference population (1000 Genomes) queried using the ldproxy function of the LDlinkR package v1.3.0^19^.

### Enrichment of MVP variants in regulatory regions

Variant enrichment was calculated as the ratio of MVP candidate variants overlapping an open chromatin region over the number of matched SNPs overlapping the same open chromatin region in a given tissue. Enrichment of MVP variants across a variety of tissues was evaluated using previously generated mitral valve tissue ATAC-Seq data (diseased mitral valve tissue from 5 individuals with MVP who underwent mitral valve replacement surgery in Göttingen Germany; non-diseased mitral valve tissue from 5 de-identified donor human hearts from Washington Regional Transplant Community in Washington, DC)^20^ as well as available bulk ATAC datasets (narrowpeaks BEDs) in all tissues from ENCODE version 2^21^. Tissue types and accession numbers are detailed in **Supplementary Table 3**. For each MVP potential functional SNP (defined as the 95% credible set plus LD proxies), we identified a set of approximately 500 matched control SNPs using the GREGOR package v1.4.0^22^. This software package generates a set of 500 control SNPs for each index SNP matching on three parameters: the number of variants in LD (e.g., if the index SNP is in LD with three variants, the matched control SNP should also have significant LD with three variants), minor allele frequency of the index SNP (+/-1%), and distance to the same nearest gene as the index SNP. Significance was determined using a one-sided binomial test (greater enrichment as the alternative hypothesis). P-values were adjusted for multiple testing by Bonferroni correction. We compared fold-enrichment between normal mitral valve, myxomatous mitral valve, other heart, and non-heart tissue samples using Wilcoxon rank-sum tests.

### Annotation of candidate variants with potential regulatory function

We evaluated whether MVP candidate variants had potential regulatory function using mitral valve bulk ATAC-seq peaks as described above and snATAC-Seq peaks from fibroblasts, (“Cardiac_Fibroblast”,”Fetal_Cardiac_Fibroblast”,”Fetal_Fibro_General_1”, and “Fibro_General” clusters), ventricular cardiomyocytes (“V_Cardiomyocyte” and “Fetal_V_Cardiomyocyte” clusters) and vascular smooth muscle cells (Vasc_Sm_Muscle_1” and “Vasc_Sm_Muscle_2” clusters) from the human enhancer atlas^23^ (tissue types and source are detailed in **Supplemental Table 3**). Variant overlap with ATAC-seq/snATAC-seq peaks was determined using the bedtools v2.29.0 annotate function. Candidate variants were annotated as being in an active promoter if they overlapped an open chromatin region and were localized to within 1500 bp of a known transcription start site (TSS). Variants were annotated as being in candidate ***cis*** regulatory elements (cCRE) if they overlapped an open chromatin region and were not localized to within 1500bp of a known TSS. Gene coordinates were retrieved from Gencode release 38. We used Integrated Genome Browser (IGB, v9.1.8) to visualize read density profiles and peak positions in the context of the human genome^24^.

### Expression quantitative trait locus (eQTL) colocalization

We performed eQTL colocalization for lead variants using MVP GWAS summary statistics and *cis*-QTL data from GTEx v8 for relevant extra-valvular tissues (fibroblasts, heart left atrial appendage, heart left ventricle). For each colocalization analysis, MVP summary data were subset to a region within 1 Mb around each lead variant, and these were merged with variant-QTL associations from tissue-specific GTEx data. Colocalization was performed using the COLOC (v5.1.0) package in R. We considered a PP4 > 0.75 as evidence of colocalization.

### Causal gene prioritization and pathway enrichment analysis

For each lead variant, we generated a list of prioritized protein-coding genes based on the following methods: 1) nearest gene, 2) genes for which MVP candidate variants were within active promoter regions, 3) closest genes from *cis-*regulatory elements overlapping with MVP candidate variants, 4) genes in promoter-enhancer loops with MVP candidate variants in cardiovascular tissues (aorta, left ventricle, or fibroblasts) as identified by Jung et al^25^, 5) extra-valvular eQTL high posterior probability colocalization, and 6) protein-altering variation, defined as a lead variant being protein-coding or in high LD (*r*^*2*^ > 0.8) with a protein-coding variant. Missense variants were retrieved from ENSEMBL SNP version 111 using biomaRt v2.54.1^26^. Coding variants were annotated as damaging if they were protein truncating or missense variants predicted by PolyPhen-2^57^ to be probably damaging or by SIFT^58^ to be deleterious.

A single causal gene was prioritized for each lead variant using the following criteria: 1) for lead variants which were either a damaging coding variant or in significant LD (*r*^*2*^ > 0.8) with a damaging protein-altering variant, the altered gene was prioritized as the most likely causal gene, 2) by consensus of the greatest number of indicators, or 3) for variants with only one indicator or with equal numbers of indicators for more than one gene, the nearest gene was prioritized. Pathway enrichment analysis for the full set of genes prioritized by any method was performed using the clusterProfiler package v4.6.2^27^.

### Gene-based association analysis

A gene-based association analysis using our MVP GWAS summary data was performed using LDAK-GBAT^28^. LDAK-GBAT assumes a linear model to test genes for association with a phenotype of interest, where the phenotypic values (vector Y) and SNP genotypes (matrix X) are considered, with each SNP genotype standardized to have a mean of zero and variance of one. LDAK-GBAT evaluates each gene’s contribution via a likelihood ratio test of the genetic variance component (σ^2^_g_), assuming genetic (σ^2^_g_) and environmental (σ^2^_e_) variance components are normally distributed. The correlation between SNPs and the phenotype, as well as SNP pairs, are calculated using summary statistics and a reference panel, allowing LDAK-GBAT to run without individual-level data.

For analysis with MVP-Gen data, we used a reference panel comprising 10,000 unrelated individuals with whole genome sequencing from the UKB^29^, with 12,832,458 SNPs (MAF ≥ 0.01) and imputation quality information (Rsq) ≥ 0.8. Gene annotations were derived from RefSeq^30^, focusing on SNPs between the transcription start and stop sites. Quality control measures were stringent, requiring a MAF ≥ 1%, SNPs to be non-ambiguous (e.g., excluding SNPs with A/T or G/C alleles which can lead to strand orientation ambiguities), and only SNPs meeting the criterion of *N* cases ≥ 0.6 × max (*N* cases) were included. After applying these filters, 6,414,352 SNPs remained, of which 2,535,072 SNPs were mapped to 17,960 genes. We used the “Human Default Model,” which sets the SNP heritability component proportional to the MAF as [MAF_j_ (1−MAF_j_)]^0.75^.

Gene-based P-values were derived using permutation-based approximations of the null distribution, and the genetic contribution for each gene was estimated. We applied Bonferroni correction to control for multiple testing across all genes, setting the significance threshold at 2.78 × 10^-6^ (= 0.05/17,960 genes). Highly correlated genes were identified using genetic contribution estimates and were clumped by excluding genes with a squared correlation > 0.1 if they shared a chromosome with a more significant gene.

### Genetic Correlations to Cardiomyopathy Trait GWAS

We compared the genetic architecture of MVP to cardiomyopathy traits by calculating genome-wide genetic correlations (rg) between our multi-ancestry MVP GWAS and the largest available summary GWAS for dilated cardiomyopathy (DCM)^31^, hypertrophic cardiomyopathy (HCM)^32^, clinical heart failure diagnosis, and its subtypes including heart failure with reduced ejection fraction (HFrEF) and heart failure with preserved ejection fraction (HFpEF)^33^. Genetic correlations were performed with LD Score Regression (LDSC), implemented using the HapMap3 CEU LD reference panel internal to LDSC.

### Collection of myocardial human tissue samples

We collected tissue from two patients with severe mitral regurgitation secondary to MVP and a clinical indication for mitral valve repair enrolled in a previously described study evaluating myocardial fibrosis in MVP^34^. Briefly, these two individuals underwent transthoracic and transesophageal echocardiography to confirm MVP and assess left ventricular function prior to surgery. Cardiac magnetic resonance (CMR) imaging using T1 mapping pre- and post-gadolinium contrast to calculate extracellular volume (ECV) fraction and assess regional tissue abnormalities, as well as late gadolinium enhancement, was performed in both patients to confirm the presence of myocardial fibrosis. Other possible causes of myocardial fibrosis including coronary artery disease, non-valvular cardiomyopathy, aortic stenosis, and previous cardiac surgery were excluded. During mitral valve repair surgery, biopsies were obtained from the inferobasal myocardium between the papillary muscles in all patients, with biopsies of the interventricular septum (IVS) to serve as within-patient controls. The study protocol was approved by the local ethics committee (study protocol number 450/18-ek at the Medical University of South Carolina) and informed written consent was obtained from each patient prior to study enrollment.

### Processing and quality control of myocardial tissue snRNA-seq data

5uM formalin-fixed paraffin-embedded (FFPE) sections were used to generate single nucleus RNA sequencing libraries using the Chromium Flex protocol (10x Genomics; Pleasanton, CA). A total of 20 slides (3-4 sections per slide) was used to isolate nuclei with a target of 400k nuclei per sample. Nuclei isolation was performed following the 10x Chromium designated protocol titled “Isolation of Cells from FFPE Tissue Sections for Chromium Fixed RNA Profiling” (CG000632). Following nuclei isolation, 10x libraries were generated following the 10x user guide for “Chromium Fixed RNA Profiling Reagent kits for singleplex samples (CG000691). Sequencing was performed by Azenta Life (Chelmsford, MA) with an expected read depth of 10,000 paired-end reads per nucleus, assuming 10,000 nuclei per sample.

Raw reads from each sample were trimmed using Skewer v0.2.2 with a minimum read length of 30 bp and a minimum quality score of 30^35^. The trimmed reads were demultiplexed, aligned, and counted using the “multi” command of 10x Genomics Cell Ranger v7.1.0. Human genome version GRCh38.p14 was used for this analysis. Empty cells were filtered out of raw cell count matrices using Cell Bender v1.0.2^36^. The Cell Bender filtered h5 count matrices were then used as inputs in the differential expression pipeline.

The results of Cell Bender were read using Seurat v5.0.3 and converted to Seurat objects^37^. The percentage of mitochondrial genes in each cell was identified by counting the number of genes containing “MT” or any variations thereof. Cells were filtered out if they contained 20% or more mitochondrial genes or contained 10,000 or more unique molecular identifier (UMI) counts. Following filtering, the data in the Seurat object was split using the SplitObject command, transformed using the SCTransform command, and integrated. We looked for integration features across 18,880 gene features prior to running the IntegrateData command. Following integration, a principal component analysis (PCA) was run on the data, corrected for batch effects using Harmony v1.2.0^38^, and clustered with a range of resolutions (from 0.1 to 1.2). We moved forward with a resolution of 0.6 for this analysis.

The clustered data was then converted to a single-cell experiment object using SingleCellExperiment v1.24.0^39^. Doublets, or instances in which multiple cells were called as a single cell, were identified by running the single-cell experiment object through ScDblFinder v1.17.0^40^. The results of ScDblFinder were assigned back to the Seurat object, and only cells classified as “singlets” were kept. The same clustering workflow described above was rerun on this filtered dataset. Each cluster was then assigned a cell-type based on gene expression. This step was performed manually by producing a dot plot of genes associated with adult human heart cells^41^ and their expression in each cluster.

### Prioritization of tissue-specific, disease-associated cell types

We evaluated the relevance of cell types in MVP risk using tissue from papillary muscle, IVS (both collected as above), and non-myxomatous mitral valves (using previously published human mitral valve single cell (sc) RNA-seq data from individuals undergoing heart transplant for non-valvular, end-stage heart failure^42^). scRNA sequencing for these latter individuals was performed using the 10x Genomics Chromium platform. scRNA-seq data in FASTQ files were processed using Cell Ranger software (version 7.1.0, 10× Genomics). Sequencing reads were aligned to the GRCh38 human reference genome and UMI counts were obtained. Subsequently, the UMI count matrix was analyzed using the Seurat package v4.3.0.1 in R v4.3.1. Cells expressing hemoglobin genes (e.g., *HBB*) were excluded as these likely represented erythrocytes. Only cells with 200–4000 detected genes and mitochondrial UMI < 10% were considered valid and included in downstream analyses. To mitigate batch effects, the Harmony method^38^ was applied to integrate MV data. After integration, principal component analysis (PCA) was conducted using the “RunPCA” function, and clustering was performed using the “FindNeighbors” and “FindClusters” functions, with a resolution of 0.3 determined by “Clustree.” Dimensionality reduction was achieved with the “RunUMAP” function.

scPagwas is a statistical method that integrates GWAS and sc/snRNA-seq to infer trait-relevant genes and cell types^43^. In brief, scPagwas calculates the correlation between the genetic effects of SNPs and pathway-specific (Kyoto Encyclopedia of Genes and Genomes [KEGG] database) gene expression levels using a linear regression model to estimate a regression coefficient τ for each cell. The resulting gene-by-cell matrix is then converted to a pathway activity score (PAS)-cell matrix, using the first principal component of pathway-specific gene expression. scPagwas then calculates the genetic correlation PAS (gPAS) for each cell and correlates it with the expression levels of each gene across all cells to prioritize trait-related genes. Finally, a trait-relevant score (TRS) is calculated for each cell by weighted summation of the expression of prioritized genes, enabling the identification of cell subpopulations that are genetically linked to specific traits. We used scPagwas to infer trait-relevant cell types using MVP-Gen GWAS summary statistics and sc/snRNA seq data from papillary muscle, septal muscle, and mitral valve tissue as described above.

### Evaluation of differential gene expression between fibrosed papillary muscle and fibrosis-free interventricular septal tissue

After isolating cell types for combined IVS and papillary muscle samples, we identified differentially expressed genes in each cell type using Seurat’s FindMarkers command^37^. We subsetted the Seurat object based on cell type; e.g., differentially expressed genes for each cell type were determined based only on results from that cell type. Samples from the IVS were treated as controls, and samples from papillary muscle were treated as the test condition. Myocardial samples (IVS versus papillary muscle) were compared for the same individual to account for potential batch effect. To ensure every gene was tested across all cell types, the minimum percentages and log-fold change thresholds were set to 0. The FindMarkers command identified differentially expressed genes using the Model-based Analysis of Single-cell Transcriptomes (MAST) v1.28.0, which employs a hurdle model: a two part generalized linear model which simultaneously considers rate of expression over the background of various transcripts and the positive expression mean^44^. Any gene with an absolute log-fold change of 0.3 or more and a Bonferroni-adjusted P-value for the number of genes tested of 0.05 or less was deemed significant. Annotations of Gene Ontology (GO) terms and Pathway enrichment were performed using EnrichR v3.2 and MSigDBR v7.5.1.9001, respectively^45,46^.

### Evaluation of mitral valve prolapse lead variants in myocardial fibrosis GWAS

To determine whether there were shared causal variants between our MVP GWAS and a recently performed GWAS of myocardial interstitial fibrosis^47^ we performed colocalization analyses for all MVP GWAS lead variants using the coloc package (v5.1.0.1) in R. We considered a window of 250 kb around each lead variant and PP_4_ ≥ 0.75 as evidence for colocalization.

## Results

### Genome-wide association study of mitral valve prolapse

We performed a multi-ancestry GWAS of MVP in autosomes among 2,257,624 individuals, representing 21,517 MVP cases. Demographic characteristics by study are presented in **Supplemental Table 1**. The resulting GWAS meta-analysis included summary data on 17,690,809 variants that were present in at least two studies and passed all quality control measures. We identified 76 independent multi-ancestry lead variants and 8 additional conditionally independent secondary signals, of which 17 were known prior MVP findings, and 67 (80%) were novel (**Figure 1, Supplemental Figure 1, Supplemental Table 4**). Of the 84 independent lead variants or secondary signals identified in multi-ancestry GWAS, three were low frequency (MAF ≥ 0.01 and < 0.05). There was no significant heterogeneity across studies for any lead variants (Q_P-value_ < 0.05/84 = 0.0006).

**Figure 1:**
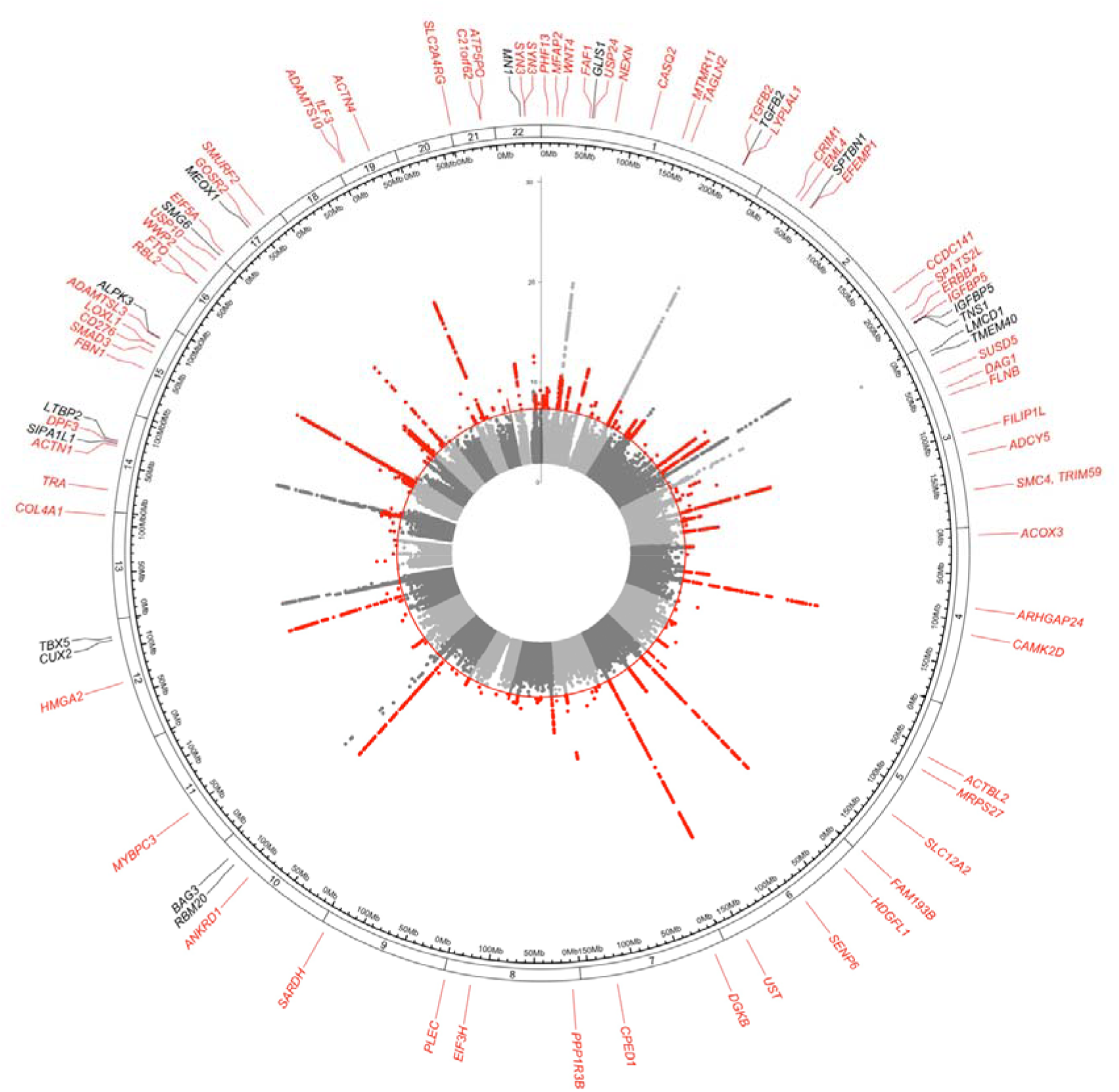
Circos plot of multi-ancestry mitral valve prolapse genome wide association study **Legend:** Circos plot for our multi-ancestry mitral valve prolapse genome-wide association study in autosomes. Prioritized genes are highlighted at corresponding chromosomal positions at the outer circle, with novel genes and SNPs in red, and known genes and SNPs in black. Genome-wide significance indicated by red circle (P-value<5×10^-8^). SNP = single nucleotide polymorphism.

In ancestry stratified populations there were 20,228 European genetic ancestry MVP cases among 2,054,365 total European genetic ancestry individuals, 946 African genetic ancestry MVP cases among 136,508 African ancestry individuals, and 343 Hispanic genetic ancestry MVP cases among 66,751 Hispanic genetic ancestry individuals. There were 77 independent European genetic ancestry lead variants, among which five were independent from any multi-ancestry MVP GWAS lead variant (**Supplemental Table 5, Supplemental Figure 2**). There were no genome-wide significant lead variants in African or Hispanic genetic ancestry GWAS (**Supplemental Figure 3, Supplemental Figure 4**). Associations by study for all primary and secondary lead variants in both our multi-ancestry and European genetic ancestry GWAS are presented in **Supplemental Table 6**.

### Prioritization of candidate causal genes and pathways

We evaluated whether MVP-associated SNPs were enriched among functionally annotated genomic regions in mitral valve (both diseased post-mitral valve replacement samples and normal mitral valve samples from deceased donors) versus other tissue types (using publicly available data from ENCODE, **Supplemental Table 3**). MVP-associated SNPs were most enriched in mitral valve tissue (both non-myxomatous and myxomatous) when compared with other heart or non-heart tissues, with a median (interquartile range) fold enrichment (relative to matched control SNPs in the same tissue) for non-myxomatous mitral valve tissue of 3.17 (3.03-3.25), myxomatous mitral valve tissue of 2.80 (2.26-2.86), compared to other heart tissues of 1.93 (1.79-1.99), and non-heart tissues of 1.63 (1.47-1.82) (**Supplemental Figure 5, Supplemental Table 7**).

To prioritize causal genes, we determined whether any potential causal variants in each significant genomic region were either missense variants, in LD with a missense variant, or had regulatory function determined using bulk mitral valve ATAC-seq data and snATAC-seq data from ventricular cardiomyocytes, fibroblasts, and smooth muscle cells. We additionally evaluated whether candidate variants were in promoter-enhancer loops using tissue from the thoracic aorta, left ventricle, and fibroblasts. There were 13 genomic regions with a missense candidate variant representing 15 unique genes (*THAP3, NEXN, CASQ2, MTMR11, EML4, MRPS27, PLEC, PARP10, SARDH, BAG3, CD276, NMB, ALPK3, ADAMTS10, SLC2A4RG*) (**Supplemental Table 8**). Of these, 5 missense variants (representing *NEXN, BAG3, CD276, ALPK3, and SLC2A4RG*) were predicted to be pathogenic by either PolyPhen-2 or SIFT. The 5 pathogenic missense variants were all common (lowest MAF = 0.10 for the A allele of rs7173476, a missense variant in *CD276*) and had modest effect estimates (maximum OR = 1.13 for rs2234962, a missense variant in *BAG3*). There were 39 genomic regions with a candidate variant (95% credible set plus LD proxies) located in an active promoter, 61 genomic regions with a candidate variant located in a cCRE, and 17 regions with candidate variants in a promoter-enhancer loop (**Supplemental Table 4**).

We performed a gene-based association analysis using LDAK-GBAT, which identified a total of 162 significant gene associations in MVP (**Supplemental Table 9, Supplemental Figure 6**). Of these, the majority (95/162 [59%]) were also represented in the list of 179 genes identified by any of our causal gene prioritization methods and 152 (94%) of genes were located within 500 kb of any MVP GWAS lead or secondary SNPs. Among the other 10 significant genes located outside of single-SNP MVP GWAS loci, the top significant genes included *HECTD4*, which encodes a ubiquitin ligase observed to be associated with left ventricular volumes^48^, and *PTPN11*, encoding a tyrosine phosphatase, mutations in which are reported in Noonan Syndrome, a genetic disorder characterized by distinctive facial features and in some cases mitral valve prolapse^49^.

We combined the above annotations (including nearest gene, whether candidate variants were missense, cCRE, in an active promotor, HiC, eQTL colocalization to relevant tissues, and whether variants were protein-coding) to prioritize a single gene for 88 significant genomic regions, and two prioritized genes for rs11924850 (**Figure 2, Supplemental Table 4**). We additionally performed pathway enrichment analysis to highlight MVP biologic pathways for the union of 246 genes identified by LDAK-GBAT or any gene prioritization method. Pathway enrichment analysis identified 75 unique significant pathways (**Supplemental Table 10**), of which the vast majority represented cardiac muscle cell processes (e.g., myofibril, sarcomere, muscle cell development).

**Figure 2:**
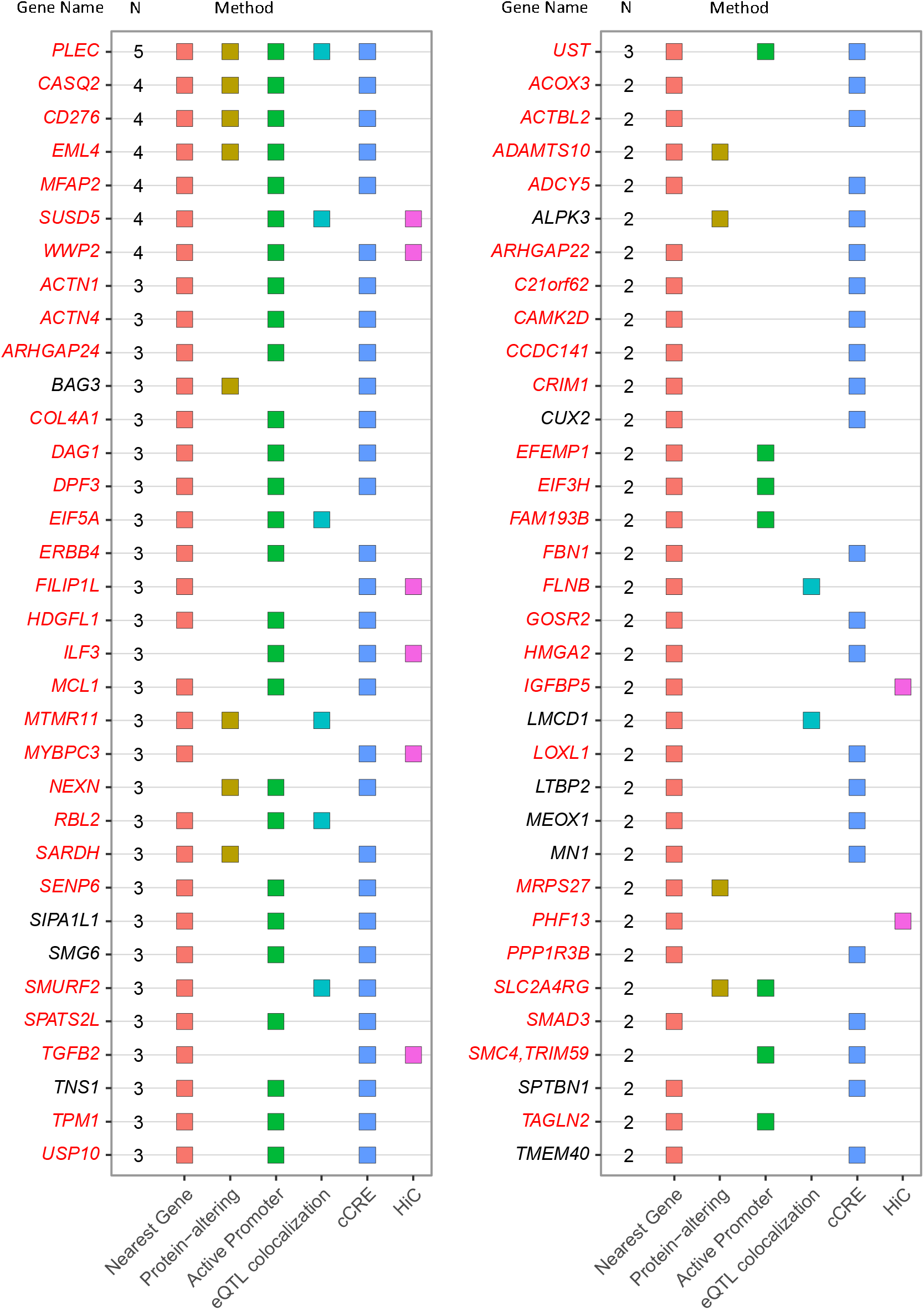
Dot plot of top prioritized genes for independent genome-wide significant mitral valve prolapse risk loci with annotation **Legend:** Dot plot indicating top prioritized gene for significant lead variants with at least two supportive methods in the mitral valve prolapse GWAS. Novel gene names are colored red, known gene names are colored black. Y-axis corresponds to functional annotations including: Nearest Gene, protein-altering variants, variants in an active promoter (Active Promoter) for a given gene, variants with significant eQTL colocalization for a given gene (eQTL colocalization), variants in candidate *cis* regulatory elements (cCRE) for a given gene, and variants which are in an enhancer loop for a given gene (HiC).

### Genetic correlations to cardiomyopathy traits

Several lead variants in MVP are also established genetic risk factors for cardiomyopathy traits, including *ALPK3* (associated with non-sarcomeric HCM^50^) and *BAG3* (associated with dilated cardiomyopathy [DCM]^51^). To assess shared genetic architecture between MVP and cardiomyopathies, we evaluated the genetic correlation between our MVP GWAS and cardiomyopathy traits including HCM, DCM, non-ischemic heart failure, as well as heart failure subtypes including heart failure with reduced ejection fraction (HFrEF) and preserved ejection fraction (HFpEF). We found a moderate, statistically significant genetic correlation between MVP and HCM, DCM, and HFrEF, with the strongest observed shared genetic architecture observed between MVP and HCM (rg = 0.38, P-value = 8.0 x 10^-19^) (**Supplemental Table 11**).

### Prioritization of disease-associated mitral valve cell types

We evaluated and compared the cellular composition of human mitral valves (characterized using previously published mitral valve scRNA-seq data^42^) and left ventricular tissue (characterized using snRNA-seq of papillary muscle and IVS samples from two individuals with MVP and mitral regurgitation requiring mitral valve repair). Mitral valve tissue exhibited a distinct cellular composition characterized by a predominance of valve interstitial cells (VICs, representing 73% of all analyzed cells), followed by myeloid cells (17.2% of analyzed cells), lymphocytes (5.8%), valvular endothelial cells (VECs, 2.2%), mast cells (0.9%), and myofibroblasts (0.9%) (**Supplemental Figure 7A**). In contrast, papillary and IVS tissue exhibited a myocardial predominant transcriptional profile primarily consisting of ventricular cardiomyocytes (papillary muscle tissue [pap]: 49.3%, IVS: 53.2%), endothelial cells (pap: 21.4%, IVS: 19.8%), and fibroblasts (pap: 15.1%, IVS: 12.1%). Additional populations included pericytes (pap: 7.4%, IVS: 6.1%), myeloid cells (2.7% in both), lymphocytes (pap: 3.0%, IVS: 2.0%), and smooth muscle cells (pap: 1.2%, IVS: 4.03%) (**Supplemental Figure 7B**,**C**). We integrated our MVP GWAS summary data with mitral valve, papillary muscle, and IVS tissue scRNA-seq data using scPagwas to identify the predominant cell types associated with risk of MVP in each tissue. In mitral valve tissue, VICs (P-value = 4.65 × 10^−3^) were the primary cell type enriched with MVP risk genes (**Supplemental Figure 7D**). In contrast, in both papillary muscle and IVS tissue, ventricular cardiomyocytes (P-value pap: 1.27 × 10^−4^, IVS: 1.95 × 10^−2^) were the most significantly enriched cell types for MVP risk genes (**Supplemental Figure 7E**,**F**).

### Characterization of genes and pathways in mitral valve prolapse related myocardial fibrosis

Individuals with MVP are prone to left ventricular fibrosis, most commonly of the inferobasal wall and papillary muscle, hypothesized as due to localized stress from abnormal valve motion^34,52^. Regional myocardial fibrosis, especially of the papillary muscles, is considered a risk factor for arrhythmia in MVP. To determine whether MVP-relevant genes are differentially expressed in myocardial fibrosis, we performed snRNA-seq of fibrotic papillary muscle and normal intraventricular septum (IVS) tissue specimens from two individuals with MVP and mitral regurgitation requiring mitral valve repair. We then compared cell-specific gene expression between the fibrosed papillary muscle tissue from these individuals with their paired normal IVS tissue. Importantly, while we would ideally compare fibrotic papillary muscle to non-fibrotic papillary controls, the latter tissue is impractical to acquire. However, the developmental origin of cell populations that give rise to the papillary muscles and left septal appendage is the same, and their molecular programs are largely conserved^53-55^.

Histological evaluation using Masson’s trichrome staining of tissue samples confirmed marked fibrosis of papillary muscle tissue specimens with substantially less fibrosis of IVS specimens (**Supplemental Figure 8**). Clustering of snRNA seq data from all samples for both individuals (papillary muscle and IVS together) revealed seven unique cell types, annotated as ventricular cardiomyocytes, fibroblasts, endothelial cells, lymphocytes, myeloid cells, smooth muscle cells, and pericytes. We compared cell type-specific gene expression between papillary muscle specimens and IVS specimens in the full set of 37,037 genes with available expression data across all seven cell types. In total, there were 4,963 genes significantly differentially expressed between papillary muscle and IVS specimens, of which 4,911 genes were differentially expressed in cardiomyocytes, 72 genes were differentially expressed in endothelial cells, 80 genes were differentially expressed in fibroblasts, two genes were differentially expressed in smooth muscle cells, and one gene was differentially expressed in pericytes (**Figure 3A, Supplemental Table 12**).

**Figure 3:**
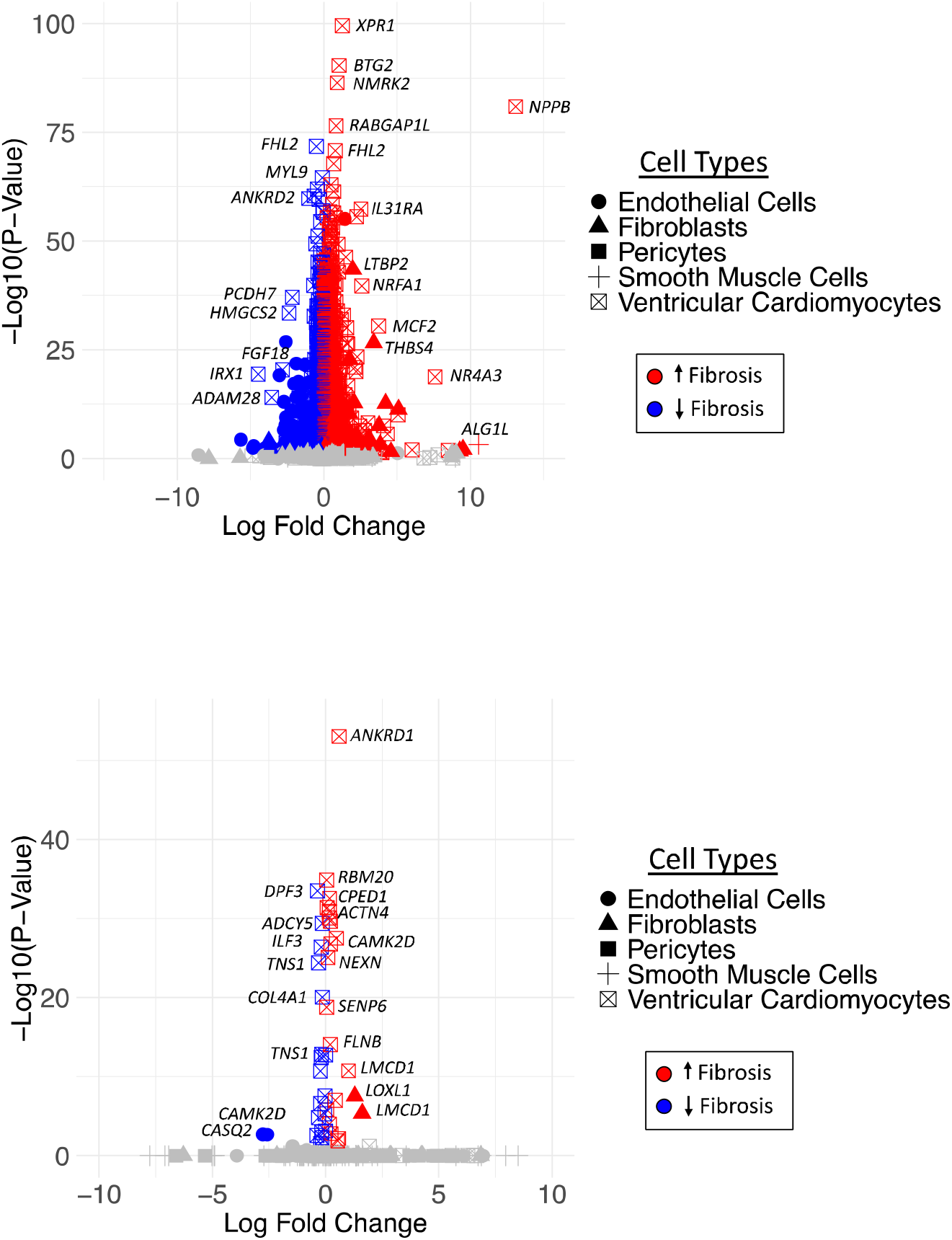
Volcano plot of differentially expressed genes by cell type comparing papillary muscle to interventricular septal tissue from individuals with severe mitral valve prolapse **Legend:** Volcano plots of differentially expressed genes comparing gene expression by cell type between papillary muscle and interventricular septum for (A) all genes and (B) genes prioritized by our mitral valve prolapse GWAS. Coloring of point estimates indicates direction of gene expression differences, with red indicating increased fibrosis and blue indicating decreased fibrosis. Grey indicates no significant difference.

Of the 4,963 genes differentially expressed between papillary muscle and IVS specimens, 50 genes (56% of 89 top prioritized genes) were also prioritized as the most likely causal gene for an MVP GWAS lead variant, which is significantly higher than what would be expected by chance (P-value < 0.001, hypergeometric test), of which the majority were differentially expressed in ventricular cardiomyocytes (**Figure 3B**). Pathway enrichment analysis of these 50 genes again highlighted a majority of pathways relevant to cardiac muscle cell processes, but additionally included cytoskeletal function (e.g., cortical actin cytoskeleton), TGF-β signaling, and the ECM (e.g., basement membrane) (**Supplemental Figure 9**).

To evaluate whether any MVP lead variants might also be a genetic risk factor for myocardial fibrosis, we performed colocalization analyses across all MVP lead genomic regions using our MVP GWAS summary statistics and summary statistics from a recently published GWAS of myocardial interstitial fibrosis^47^. Of the 89 MVP lead variants, one variant, rs17046126 (intronic to *CAMK2D*) was considered likely to be causal for both MVP and myocardial interstitial fibrosis (**Supplemental Table 13, Supplemental Figure 10**). This variant was sub-genome-wide significant in GWAS for myocardial interstitial fibrosis (rs17046126, OR [95% CI] for MVP risk allele T: 1.04 [1.02-1.05], P=3.4 × 10^-7^) and had a concordant direction of effect (the risk allele for MVP also increased risk of myocardial fibrosis). The top prioritized gene for this variant, *CAMK2D*, was also significantly differentially expressed between fibrosed papillary muscle and normal IVS tissue. We additionally interrogated the summary statistics of a recent GWAS of electrocardiogram (ECG) traits in the UKB^56^, finding that rs17046126 has genome-wide significant associations with ECG traits in the early myocardial repolarization period (**Supplemental Figure 11**).

## Discussion

We performed a GWAS of MVP encompassing 21,517 cases among a total sample of over 2.2 million individuals, identifying 89 genomic risk loci for MVP, of which 72 were novel findings. We prioritized causal genes and pathways using mitral valve and extra-valvular epigenetic and transcriptomic data, identifying candidate variants with coding or regulatory function in a majority of our significant genomic regions. We performed genetic correlations between MVP and cardiomyopathy traits, identifying overlap in the genetic architecture of HCM and MVP. Finally, we interrogated scRNA-seq data in human cardiac muscle tissue from individuals with severe MVP and intersected results with publicly available genomics data on myocardial interstitial fibrosis to characterize genes associated with risk of papillary muscle fibrosis, identifying a gene, *CAMK2D*, with evidence for a shared role in both myocardial fibrosis and MVP.

Our data permit several relevant conclusions. First, we confirm that genetic risk of MVP is driven predominantly by mitral valve-specific effects, predominantly via valve interstitial cells (VICs), and again establish a role for the extracellular matrix (ECM) in the pathobiology of MVP. Evaluation of MVP GWAS SNPs among chromatin accessible genomic regions across tissue types clarified that genetic risk loci for MVP were most enriched in open chromatin regions in the mitral valve, indicating that these loci likely modify mitral valve-specific gene expression. Integration of scRNA-seq mitral valve data with MVP GWAS results revealed that VICs constitute the predominant MVP-associated cell type in mitral valve tissue. This is consistent with histopathologic and *in vitro* studies demonstrating that VICs transition to an activated form in MVP which can promote ECM remodeling^1,57^. Many of the top prioritized genes from MVP lead variants encode ECM proteins or regulate ECM-relevant pathways. These include replicated MVP genetic risk factors from prior GWAS such as *TGFB2* and *LTBP2*, but also novel risk genes in MVP such as *COL4A1*, encoding type IV collagen, which is a component of the basement membrane, *ADAMTS10*, encoding an ECM protease, and *LOXL1*, encoding an enzyme required for collagen cross-linking.

Second, we extend evidence for shared genetic risk between cardiomyopathy and MVP. In a prior MVP GWAS, authors highlighted significant associations between MVP and variants in *ALPK3, BAG3*, and *RBM20*^*10*^. These genes are known to contribute to both hypertrophic and dilated cardiomyopathy^50,58,59^. In the current GWAS, we replicate these findings and identify several additional cardiomyopathy-relevant genes, particularly genes pleiotropic to HCM, including MVP lead variants prioritized to *NEXN*, encoding the cardiac Z-disc protein nexilin^60^, and *MYBPC3*, encoding myosin-binding protein C^61^. Accordingly, we found a modest, statistically significant genetic correlation between MVP and HCM, suggesting shared genetic architecture between these two conditions. Histopathological evaluation of mitral valves from individuals with HCM feature thickened and elongated valve architecture, similar to what is observed in MVP, suggesting that there may be a unifying underlying biologic process^62^. Further research is necessary to determine whether these shared pathways represent underlying shared biology or confounding due to the presence of co-occuring or interacting conditions.

Finally, we analyzed transcriptomic data from human tissue samples in individuals with MVP and myocardial fibrosis, an established risk factor for arrhythmic MVP, to identify MVP genetic risk factors that may also contribute to cardiac fibrosis. Interestingly, approximately half of the top prioritized genes identified in our MVP GWAS were significantly differentially expressed between fibrosed papillary muscle and fibrosis-free IVS. A variant in one of these genes, *CAMK2D*, was near genome-wide significant in a recent GWAS of myocardial interstitial fibrosis and is suggested to be the causal variant in both our MVP GWAS and the myocardial interstitial fibrosis GWAS in colocalization analysis. This variant was also observed to be associated with ECG changes in early myocardial repolarization, represented by the early ST segment interval, changes that are also associated with risk of arrhythmic events among individuals with MVP^63^. *CAMK2D* encodes a calmodulin-dependent protein kinase involved in calcium ion transport that has been previously identified as a GWS signal in GWAS for atrial fibrillation^64^, bradyarrhythmia^65^, and non-ischemic all cause heart failure^33^. Overactivation of CAMK2D signaling is thought to result in increased inflammation, apoptosis, and fibrosis, all mechanisms which might plausibly both increase risk for progressive degenerative mitral valve disease as well as accompanying myocardial tissue fibrosis and future risk of arrhythmia^66^.

There are several limitations to the current work. First, while we attempted to primarily use an echocardiographic diagnosis for MVP, some contributing studies relied solely or partially on a claims-based diagnosis. The accuracy of claims data for diagnosis of MVP is unknown. Furthermore, MVP is characterized histologically as two distinct forms, Barlow disease with myxomatous degeneration, and fibroelastic deficiency. We were unable to distinguish between these phenotypes, which may have different genetic risk factors. Additionally, while the present study includes substantially more individuals of non-European ancestry than prior efforts (203,295 non-European ancestry individuals compared to none in prior studies), the bulk of the dataset (91% of total study population) is of European ancestry. Finally, our evaluation of genes differentially expressed in myocardial tissue comparing regions with and without fibrosis is limited to two individuals due to the practical challenges in acquiring relevant human tissue samples, which potentially limits generalizability of results. Additionally, we cannot preclude that gene expression differences observed between fibrosed papillary muscle versus fibrosis-free IVS are due to differences in gene expression between tissue.

In summary, we performed the largest GWAS of MVP to date, identifying 72 novel genetic risk factors for the disease. Genomic and transcriptomic data again highlight genes involved in TGF-β signaling and ECM biology as disease associated mechanisms in MVP. We additionally identify cardiomyocyte biology as an important genetically mediated biologic pathway in MVP and identify a significant genetic correlation between MVP and HCM. Finally, we highlight evidence for a shared causal gene, *CAMK2D*, associated with risk of both MVP and myocardial fibrosis, which is recognized to increase the risk of malignant arrhythmia in MVP.

## Data availability

Genome-wide summary statistics generated in this study will be deposited in the CVD knowledge portal (https://cvd.hugeamp.org/) and will be accessible under accession number [to be added upon acceptance].

