## Supplementary text and figures for "Genetic Architecture and Myocardial Fibrotic Remodeling in Mitral Valve Prolapse"

**Table of Contents:**

I. **Supplemental Methods**
   a. **Study Descriptions**
II. **Supplemental Figures**
    a. **Supplemental Figure 1:** Manhattan plot and QQ plot of multi-ancestry MVP GWAS
    b. **Supplemental Figure 2:** Manhattan plot and QQ plot of European genetic ancestry MVP GWAS
    c. **Supplemental Figure 3:** Manhattan plot and QQ plot of African genetic ancestry MVP GWAS
    d. **Supplemental Figure 4:** Manhattan plot and QQ plot of Hispanic genetic ancestry MVP GWAS
    e. **Supplemental Figure 5:** Enrichment of mitral valve prolapse associated single nucleotide polymorphisms in open chromatin regions from bulk ATAC (including valve and all ENCODE tissues)
    f. **Supplemental Figure 6:** Miami plot of LDAK-GBAT results
    g. **Supplemental Figure 7:** single cell characterization and determination of disease-associated cell types of mitral valve, papillary muscle, and interventricular septal tissue
    h. **Supplemental Figure 8:** Histological evaluation of papillary and interventricular septal tissue
    i. **Supplemental Figure 9:** Gene ontology concept network plot for 44 genes prioritized by mitral valve prolapse GWAS and differentially expressed between fibrosis and no-fibrosis samples
    j. **Supplemental Figure 10:** P-P plot of MVP GWAS and myocardial fibrosis GWAS
III. **Author Funding, Disclosures, and Acknowledgements**

**Study Descriptions:**

*BioMe*

The Mount Sinai Bio*Me* Biobank is an ongoing electronic health record-linked biorepository that enrolls participants non-selectively from the Mount Sinai Health System, comprising approximately 60,000 participants. Genotyping was performed using the Global Screen Array (GSA), for ~40,000 samples, and the Global Diversity Array (GDA), for ~20,000 samples and following standard genotyping quality control. The genotyped data were imputed using the TOPMed imputation server (version r2). Mitral valve prolapse case/control status was determined using either a single instance of the *ICD-10* code I34.1 or echocardiographic diagnosis of MVP. A GWAS for MVP was performed in autosomes separately by genetic ancestry using imputed data from 524 European genetic ancestry cases, 18,212 European genetic ancestry controls, 140 Hispanic genetic ancestry cases, and 15,835 Hispanic genetic ancestry controls, 131 African genetic ancestry cases, and 14,397 African genetic ancestry controls. The associations were modelled using logistic regression in REGENIE with adjustment for age, sex, genotyping chip, and ancestry-specific principal components.

*BioVU*

BioVU (Vanderbilt University) is Vanderbilt's biorepository of DNA and genetic data extracted from discarded blood collected during routine clinical testing and linked to de-identified medical records. BioVU contains approximately 318,000 participants. Genotyping was performed using the Infinium Expanded Multi-Ethnic Genotyping Array (MEGA^EX^), and following standard genotyping quality control, the genotyped data were imputed using the TOPMed imputation server (version r2). Mitral valve prolapse case/control status was determined using either a single instance of the *ICD-10* code I34.1 or echocardiographic diagnosis of MVP. A GWAS for MVP was performed in autosomes separately by genetic ancestry using imputed data from 444 European genetic ancestry cases and 73,409 European genetic ancestry controls. The associations were modelled using logistic regression in SAIGE with adjustment for age, sex, and ancestry-specific principal components.

*Colorado Center for Precision Medicine*

The Colorado Center for Precision Medicine (CCPM) is a biobank developed by the University of Colorado Anschutz Medical Campus and UCHealth which currently has more than 200,000 participants and over 33,000 genotyped individuals. Genotyping was performed using the Illumina MEGA/exome array and following standard genotyping quality control, the genotyped data were imputed using TOPMed (version r2). Mitral valve prolapse case/control status was determined using either a single instance of the *ICD-10* code I34.1 or echocardiographic diagnosis of MVP. A GWAS for MVP was performed in autosomes using imputed data from 732 European genetic ancestry cases and 71,769 European genetic ancestry controls. The associations were modeled using logistic regression in REGENIE (v3.2.1) with adjustment for age, sex, and ancestry-specific principal components.

*Copenhagen Hospital Biobank and the Danish Blood Donor Study*

The Copenhagen Hospital Biobank (CHB) was started in 2009 and is a collection of surplus material from diagnostic testing on patients admitted to hospitals in the Danish Capital. The current analysis was done under the CHB Cardiovascular Disease Cohort (CHB-CVDC) (approval number: NVK-1708829, P-2019-93). The analysis was combined with data on participants in The Danish Blood Donor Study (DBDS) (approval number: NVK-1700407, P-2019-99). DBDS has since 2010 included blood donors into a prospective cohort study and biobank. Genotyping of both CHB-CVDC and DBDS was performed at deCODE genetics using the Illumina Global Screening Array and following standard genotyping quality control, the genotyped data were imputed using a reference panel from whole-genome sequencing of 15,576 individuals from Scandinavia, the Netherlands, and Ireland at deCODE genetics. Mitral valve prolapse case/control status was determined using either a single instance of the *ICD-10* code I34.1 or echocardiographic diagnosis of MVP. A GWAS for MVP was performed in autosomes using imputed data from 1,161 European genetic ancestry cases and 70,521 European genetic ancestry controls. The associations were modeled using logistic regression in REGENIE with adjustment for age, sex, and ancestry-specific principal components.

*deCODE*

The deCODE dataset included Icelandic patients diagnosed with MVP in the years 1983-2019 at Landspitali – The National University Hospital in Reykjavik, the only tertiary referral center in Iceland. Following long-range phasing, variants identified in 63,460 whole genome sequenced (WGS) Icelanders were imputed into 173,025 individuals chip-genotyped employing multiple Illumina platforms. Familial imputation of genotypes in first- and second-degree relatives was used to increase sample size. Mitral valve prolapse case/control status was determined using a single instance of the *ICD-10* code I34.1. A GWAS for MVP was performed in autosomes using imputed data from 738 Icelandic cases and 330,674 Icelandic controls. The associations were tested using logistic regression under the additive model, including current age or age at death, sex, and county of birth as covariates. For the association analyses, we used software developed at deCODE genetics. We used LD score regression intercepts to adjust the χ2 statistics and avoid inflation due cryptic relatedness and stratification, using a set of 1.1 million variants. P values were calculated from the adjusted χ2 results.

*FinnGen*

The FinnGen Study is an ongoing prospective cohort study that combined population-based legacy cohorts, disease-based cohorts, and individuals recruited by biobanks. FinnGen contains approximately 500,000 participants recruited in university hospital settings. Genotyping was performed using the custom Axion FinnGen1 array, as well as several non-custom legacy chips, and following standard genotyping quality control, the genotyped data were imputed using the population-specific Sequencing Initiative Suomi (SISu) v.3 imputation reference panel. Mitral valve prolapse case/control status was determined using a single instance of the *ICD-10* code I34.1. A GWAS for MVP was performed in autosomes using imputed data from 4,173 European genetic ancestry cases and 448,327 European genetic ancestry controls. The associations were modelled using logistic regression in REGENIE with adjustment for age, sex, and ancestry specific principal components.

*HerediGene (Intermountain)*

The HerediGene Population Study is a collaboration between deCODE genetics and Intermountain Health in Utah which recruits individuals in the Intermountain Health system, encompassing 22 hospitals in the Mountain West region of Utah and Idaho. Sequence variants, identified through WGS of 23,288 Americans of European ancestry from Utah, were imputed into 138,006 individuals enrolled at multiple Intermountain Healthcare facilities and chip-typed at deCODE genetics using Illumina platform. The imputation was based on long-range phasing and sequence variant calling that was performed jointly for several sample-sets (Danish, N-American, Iranian, Swedish, and Dutch), for which 50,839 have been WGS and 1,041,174 chip-typed. The associations were tested using logistic regression under the additive model, including current age or age at death, sex, and ancestry-specific principal components as covariates. For the association analyses, we used software developed at deCODE genetics. We used LD score regression intercepts to adjust the χ2 statistics and avoid inflation due cryptic relatedness and stratification, using a set of 1.1 million variants. P values were calculated from the adjusted χ2 results. Mitral valve prolapse case/control status was determined using a single instance of the *ICD-10* code I34.1. A GWAS for MVP was performed in autosomes using imputed data from 508 European genetic ancestry cases and 92,668 European genetic ancestry controls. Participants in the HerediGene population study are voluntary US residents over the age of 18 years, who gave permission to link anonymized genotypic data with electronic health records.

*Million Veteran Program*

The Million Veteran Program is an observational cohort study and large biobank in the Department of Veterans Affairs VA Healthcare System, with enrollment begun in 2011. Genotyping was performed using a custom Axiom array (MVP1.0), and following standard genotyping quality control, the genotyped data were imputed using the TOPMed imputation server (version r2). Mitral valve prolapse case/control status was determined using either a single instance of the *ICD-10* code I34.1 or regular expression query evidence for mitral valve prolapse diagnosis in clinical note text. A GWAS for MVP was performed in autosomes separately by genetic ancestry using imputed data from 4,618 European genetic ancestry cases, 459,093 European genetic ancestry controls, 203 Hispanic genetic ancestry cases, and 50,573 Hispanic genetic ancestry controls, 815 African genetic ancestry cases and 121,165 African ancestry controls. The associations were modeled using logistic regression in REGENIE with adjustment for age, sex, and ancestry-specific principal components.

*Penn Medicine Biobank*

The Penn Medicine Biobank is an academic biobank at the University of Pennsylvania healthcare system. Genotyping was performed using the Illumina Global Screening Array v.2.0, and following standard genotyping quality control, the genotyped data were imputed using the TOPMed imputation server (version r2). Mitral valve prolapse case/control status was determined using either a single instance of the *ICD-10* code I34.1 or echocardiographic diagnosis of MVP. A GWAS for MVP was performed in autosomes using imputed data from 407 European genetic ancestry cases and 26,284 European genetic ancestry controls. The associations were modeled using logistic regression in SAIGE with adjustment for age, sex, and ancestry-specific principal components.

*Munich MVP Cohort*

The Munich MVP cohort comprised patients from the KaBi-DHM biobank who were diagnosed with mitral valve prolapse (MVP) at the Department of Cardiovascular Surgery or the Department of Cardiology, TUM University Hospital – German Heart Center. Eligible patients had either undergone mitral valve surgery or presented with MVP and mitral regurgitation between March 2002 and January 2021. MVP diagnosis was confirmed using surgical reports and / or echocardiographic data. Genotyping of cases was performed with the Infinium Global Screening Array (GSA MD v3 and Control Dashboard; Illumina, San Diego, CA, USA). The control group consisted of European participants from the population-based PROCAM-2 study cohort^1^. The PROCAM-2 Study was initiated and conducted by the Leibniz Institute for Arteriosclerosis Research at the University of Münster under the leadership of G. Assmann. After his retirement, all data were transferred to the university for further scientific use. The later follow-up assessment was carried out with funds from the Institute of Epidemiology and Social Medicine, and DNA isolation was performed with financial support from the Dean of the medical faculty, both at the University of Münster. Genotyping was enabled through funds from the German Center for Cardiovascular Disease (DZHK) and was conducted using the Illumina Global Screening Array (GSA) v1.0. Phenotype and genotype data were collected after obtaining informed consent from all participants. The study was approved by the local Ethics Committee of the Technical University of Munich and the University of Münster. Imputation of 4.8 million common SNPs (MAF <0.01 in 1000 Genomes Project Europeans, phase 3 haplotype reference panel), was carried out using IMPUTE (v2). GWAS was performed with the software PLINK (Version 1.9 and 2.0). Linear regression was applied for continuous traits, while logistic regression was used for binary traits, with covariates including age, sex, principal components (the first ten to account for population stratification). To adjust for population stratification, multidimensional scaling was performed in PLINK.

**Supplemental Figure 1:** Manhattan plot and QQ plot of multi-ancestry MVP GWAS

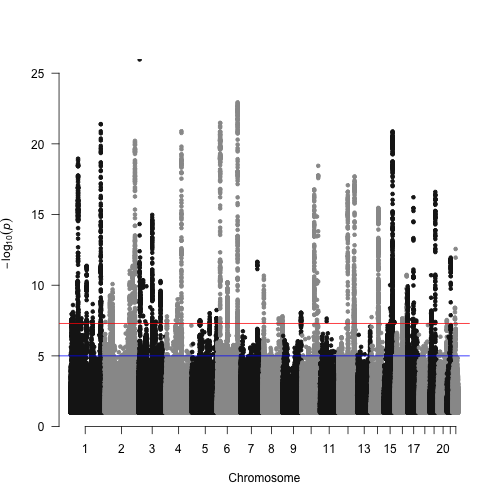
**A: Manhattan plot**

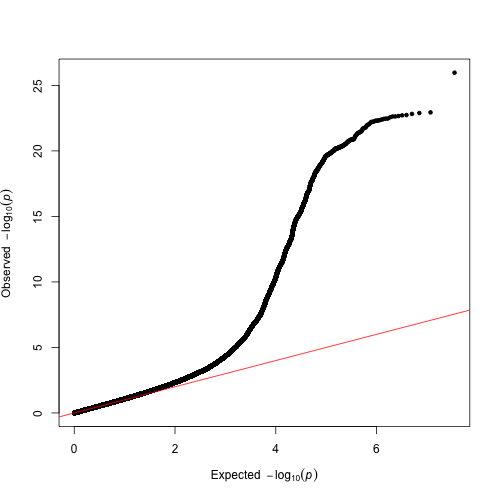
**B: QQ plot**

**Legend:** Manhattan (A) and QQ (B) plots for multi-ancestry genome wide association study in autosomes (N = 2,257,624). Genome-wide significance (P-value = 5 × 10^-8^) indicated by a horizontal red line. Abbreviations as follows, -log_10_(*p)*: negative logarithm base 10 of p-value.

**Supplemental Figure 2:** Manhattan plot and QQ plot of European genetic ancestry MVP GWAS

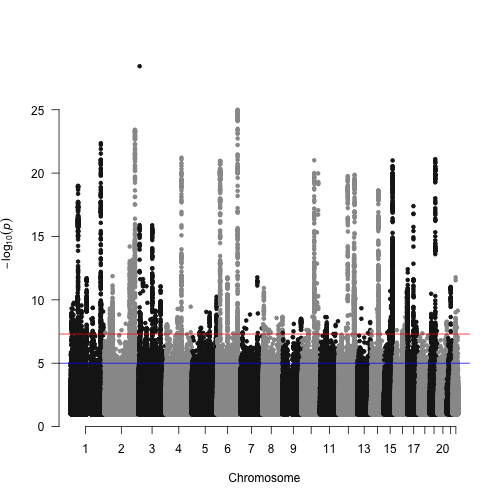
**A: Manhattan plot**

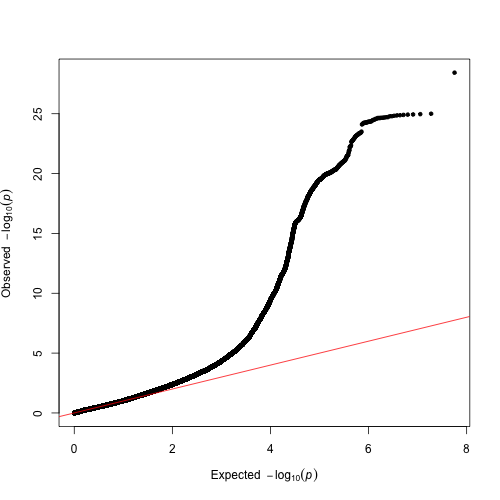
**B: QQ plot**

**Legend:** Manhattan (A) and QQ (B) plots for European ancestry genome wide association study in autosomes (N = 2,054,365). Genome-wide significance (P-value = 5 × 10^-8^) indicated by a horizontal red line. Abbreviations as follows, -log_10_(*p)*: negative logarithm base 10 of p-value.

**Supplemental Figure 3:** Manhattan plot and QQ plot of African genetic ancestry MVP GWAS

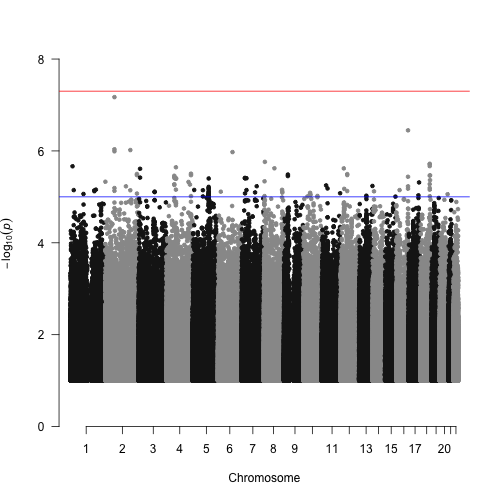
**A: Manhattan plot**

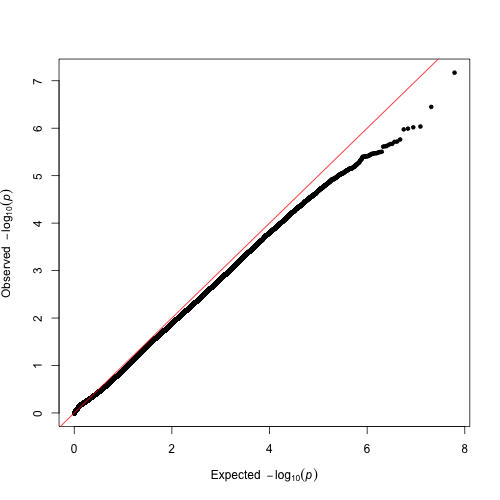
**B: QQ plot**

**Legend:** Manhattan (A) and QQ (B) plots for African ancestry genome wide association study in autosomes (N = 136,508). Genome-wide significance (P-value = 5 × 10^-8^) indicated by a horizontal red line. Abbreviations as follows, -log_10_(*p)*: negative logarithm base 10 of p-value.

**Supplemental Figure 4:** Manhattan plot and QQ plot of Hispanic genetic ancestry MVP GWAS

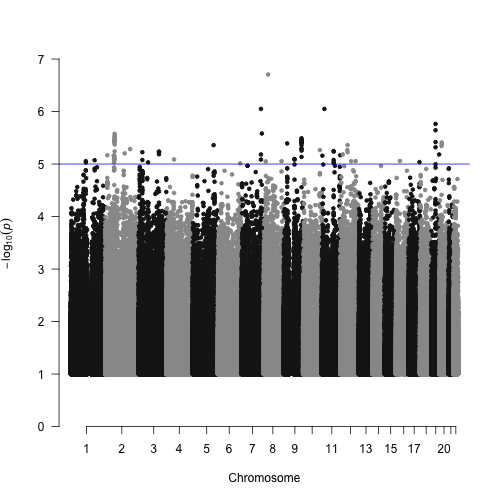
**A: Manhattan plot**

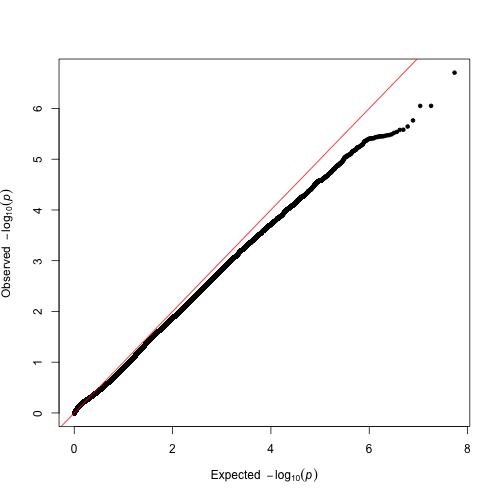
**B: QQ plot**

**Legend:** Manhattan (A) and QQ (B) plots for Hispanic ancestry genome wide association study in autosomes (N = 66,751). Genome-wide significance (P-value = 5 × 10^-8^) indicated by a horizontal red line. Abbreviations as follows, -log_10_(*p)*: negative logarithm base 10 of p-value.

**Supplemental Figure 5:** Enrichment of mitral valve prolapse associated single nucleotide polymorphisms in open chromatin regions from bulk ATAC (including valve and all ENCODE tissues).

A) Box plots of fold enrichment by tissue type

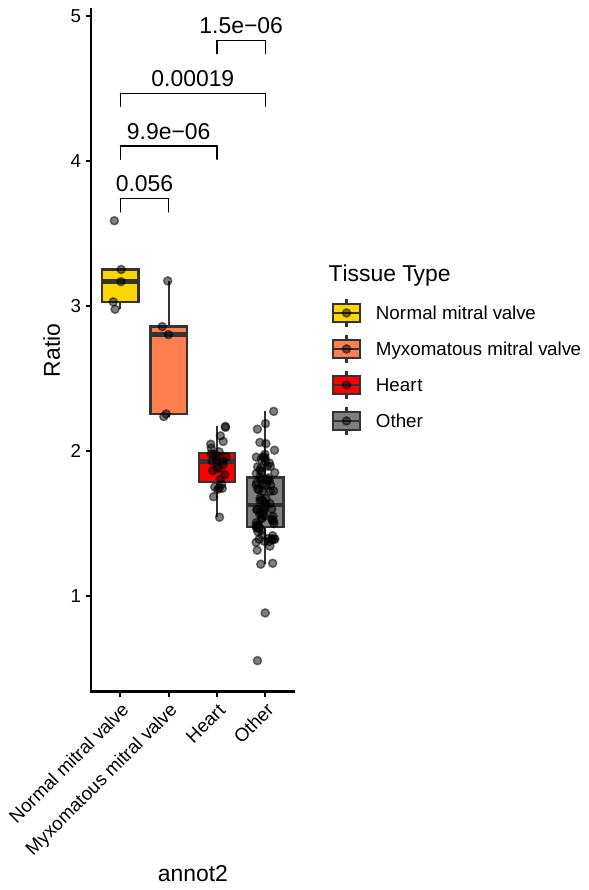

Fold-Enrichment

B) Scatter plot of P-values (Y-axis) by fold-enrichment (X-axis)

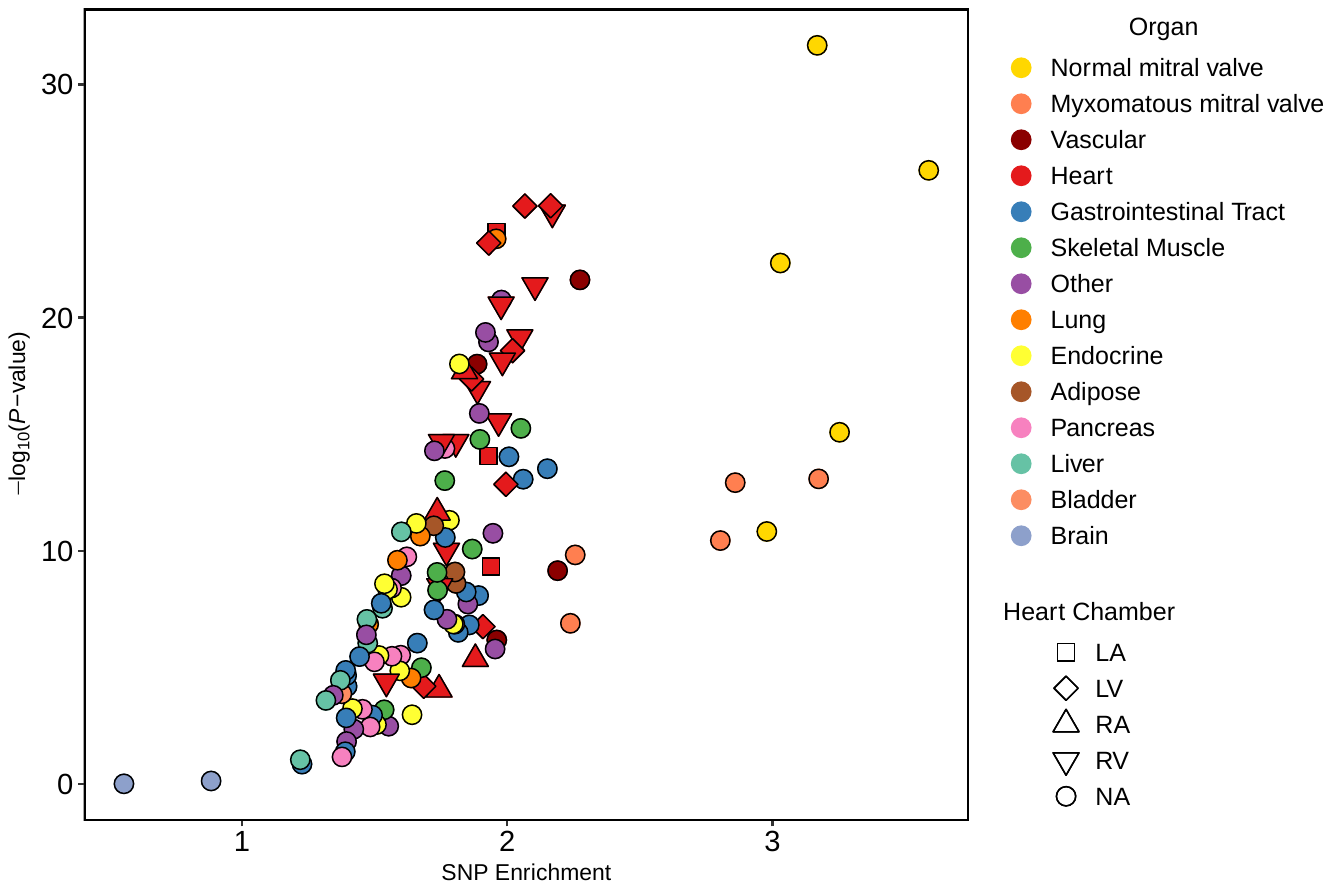

**Legend:** Box plots (A) demonstrating the median and interquartile range for fold-enrichment (Y-axis) of MVP lead SNPs compared to matched control SNPs in normal mitral valve, myxomatous mitral valve, heart, or other tissue types (see Supplemental Table 3 for a list of tissues). P-values correspond to Wilcoxon rank sum tests for difference between groups. Scatter plot (B) showing log_10_(P-value) on the Y-axis and fold enrichment of single nucleotide polymorphisms (SNPs) on the X-axis for mitral valve prolapse risk loci using bulk ATAC-seq data from various tissue types. Abbreviations as follows: SNP (single nucleotide polymorphism); LA (left atrium); LV (left ventricle); RA (right atrium); RV (right ventricle)

**Supplemental Figure 6:** Miami plot of LDAK-GBAT and multi-ancestry MVP GWAS results

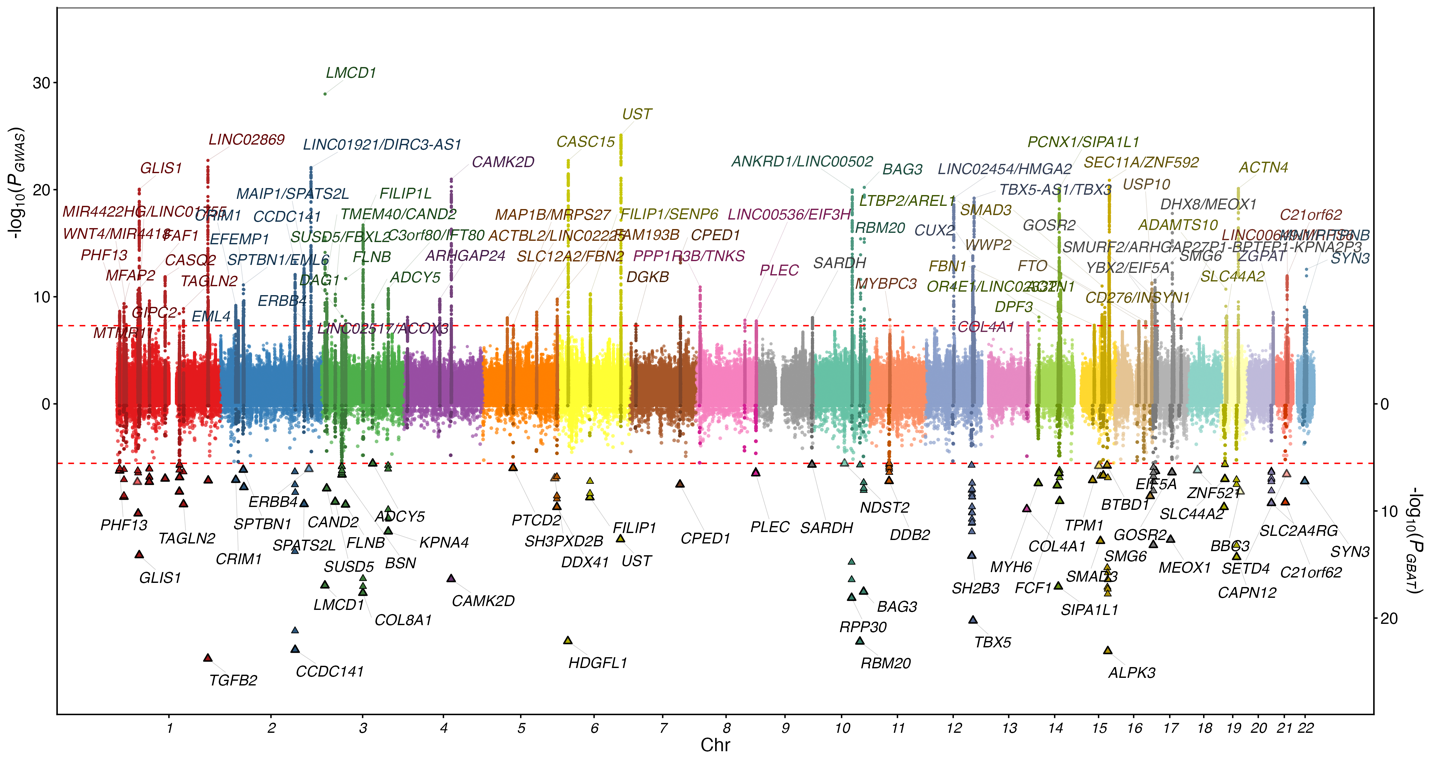

**Legend:** Miami plot with multi-ancestry genome-wide association study of mitral valve prolapse (top), labeled by nearest gene and LDAK-GBAT results (bottom). Y-axis corresponds to -log_10_(p-value).

**Supplemental Figure 7:** Single cell characterization and determination of disease-associated cell types of mitral valve, papillary muscle, and interventricular septal tissue

A)
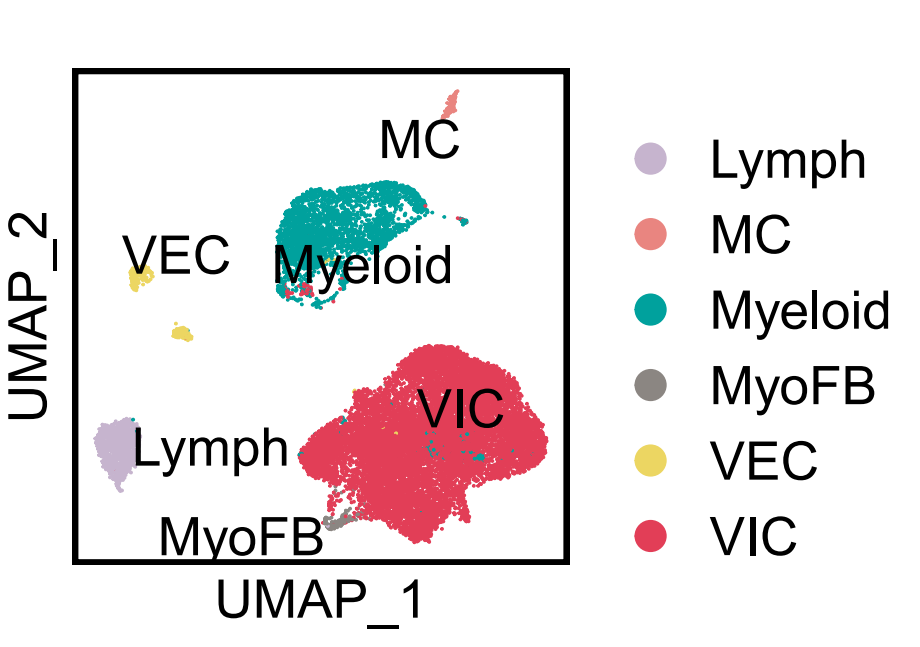
Mitral valve tissue UMAP (left) and pie chart of cell types (right)

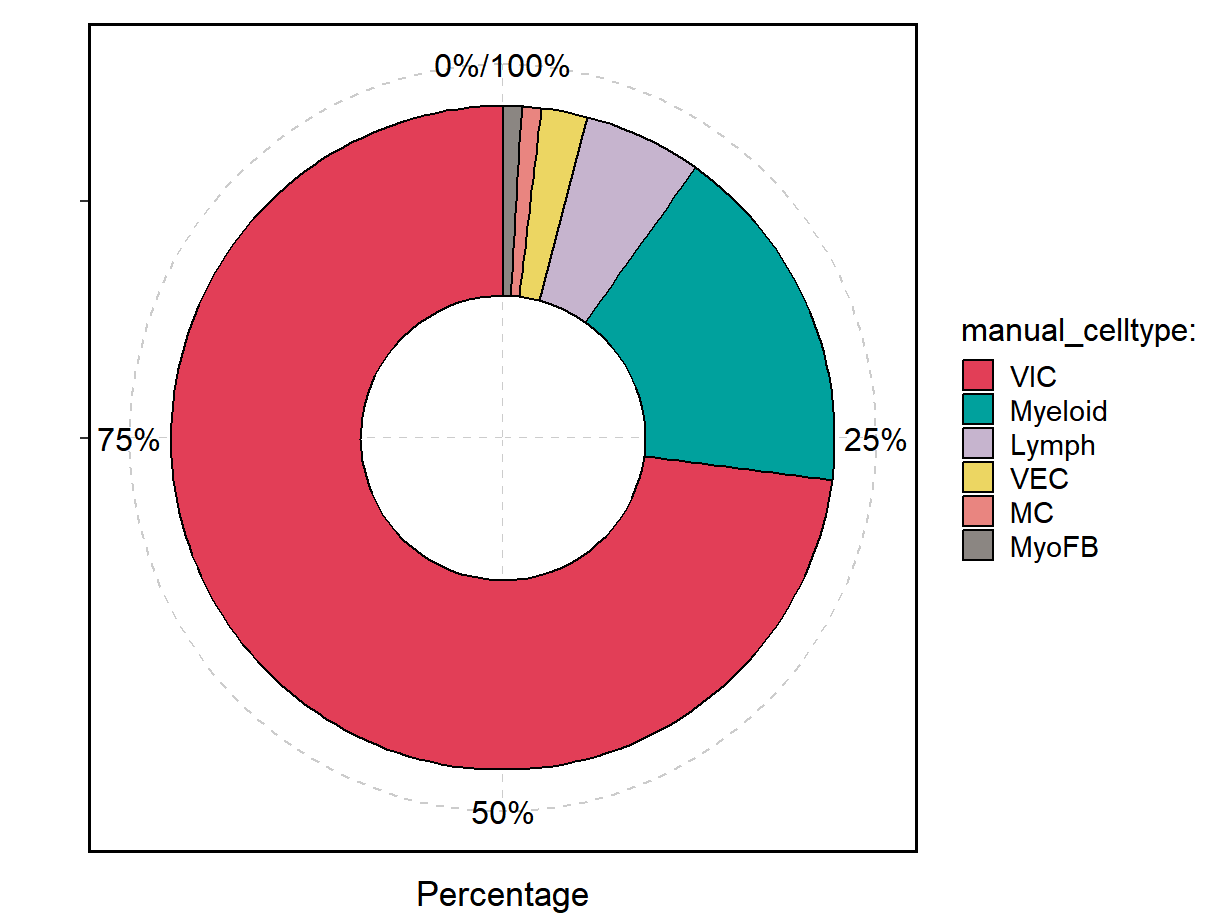

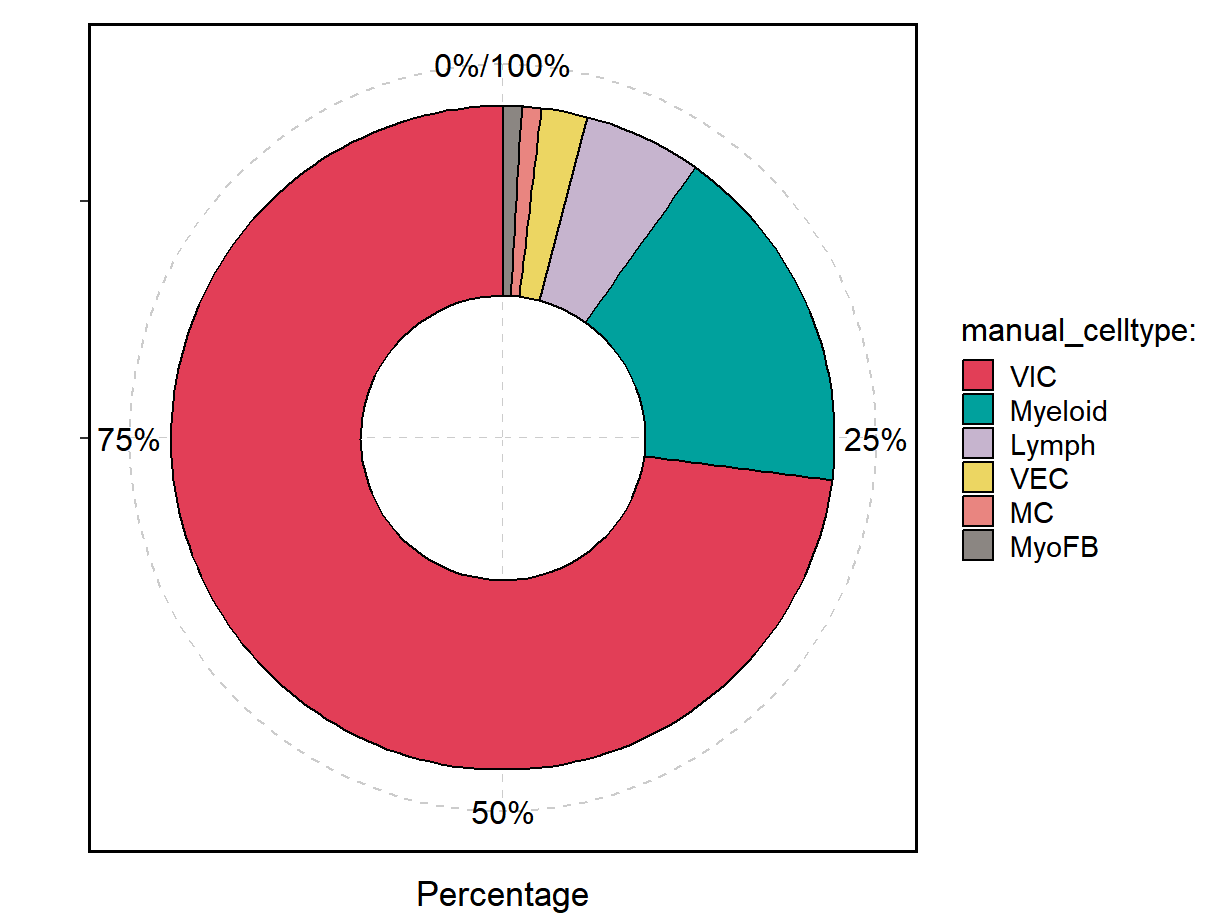

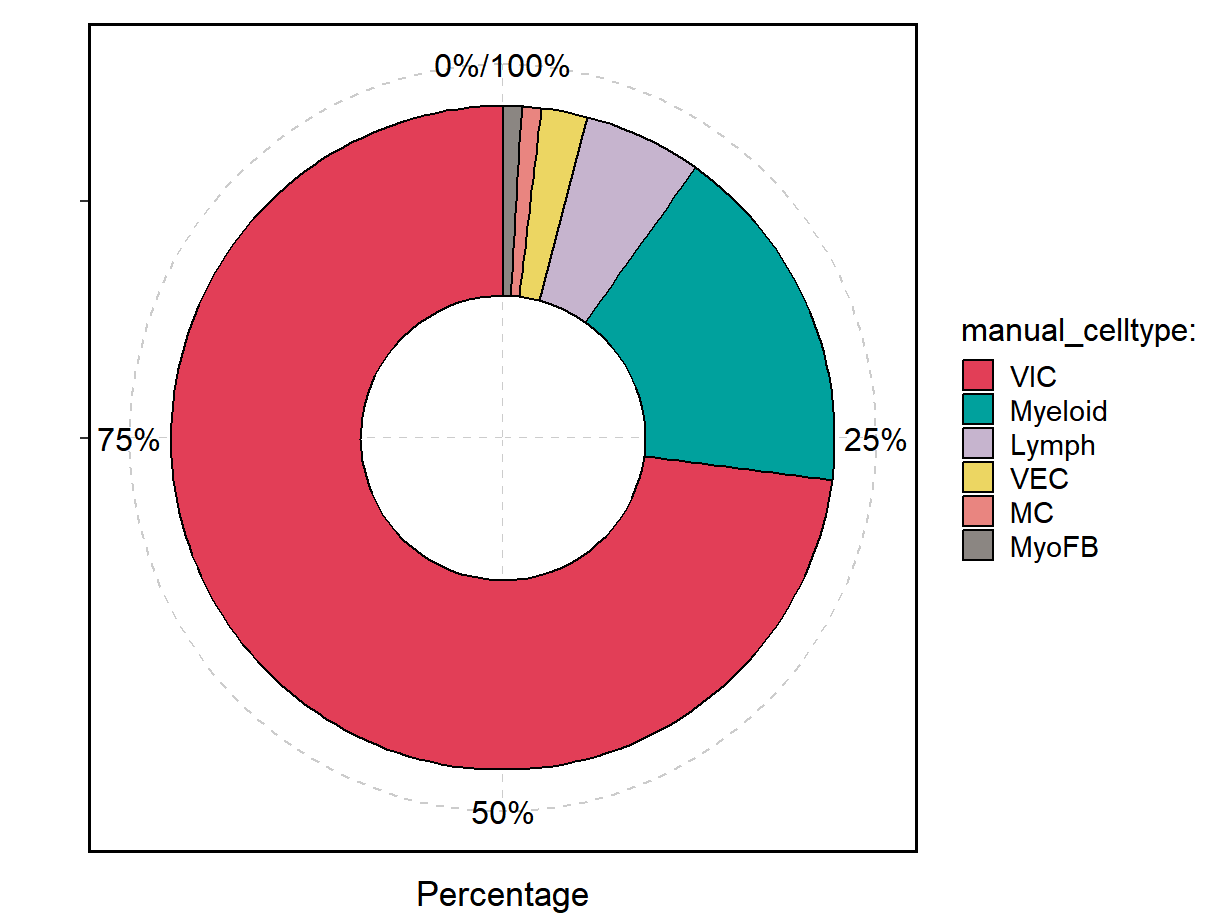

B)
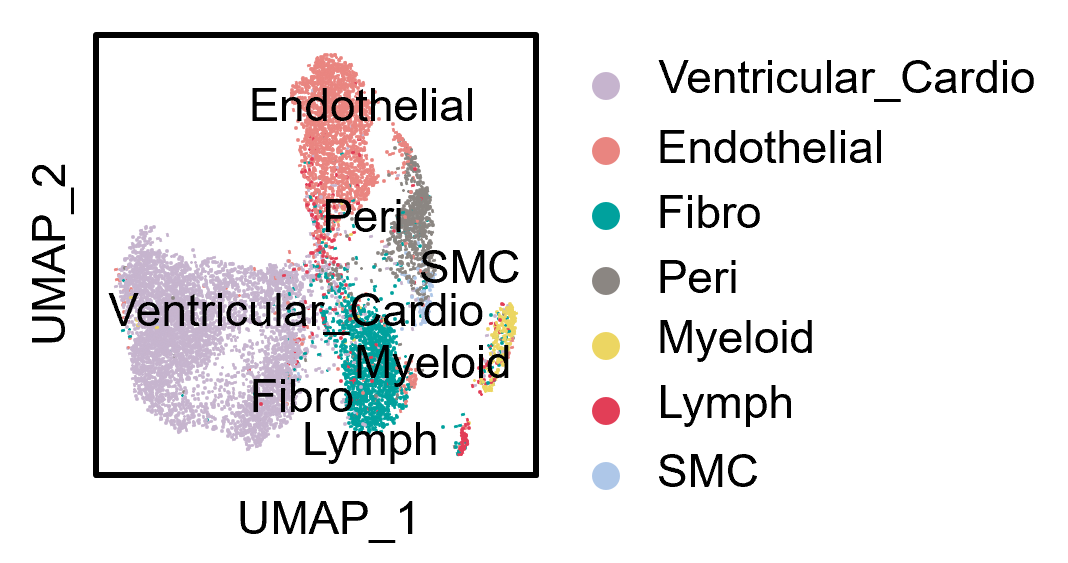
Papillary muscle UMAP (left) and pie chart of cell types (right)

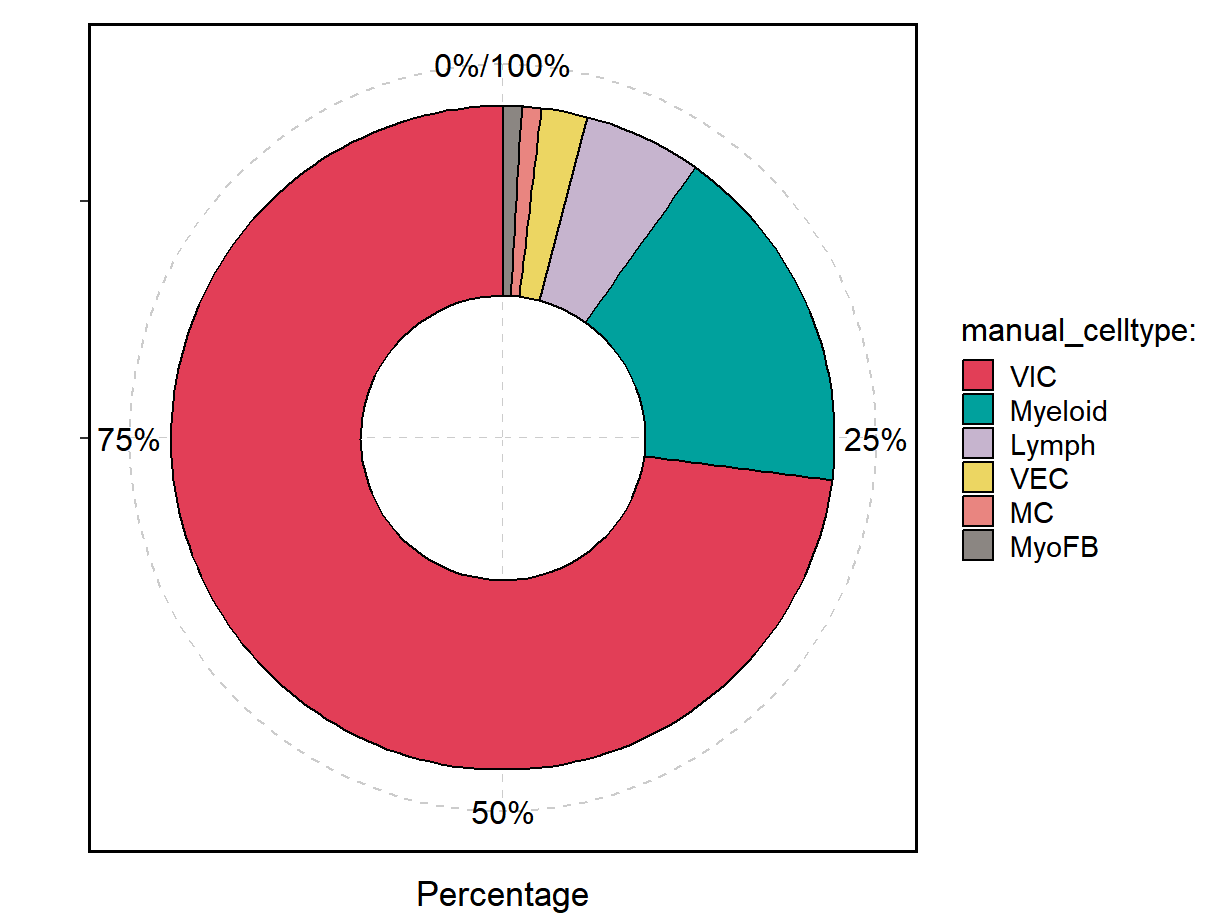

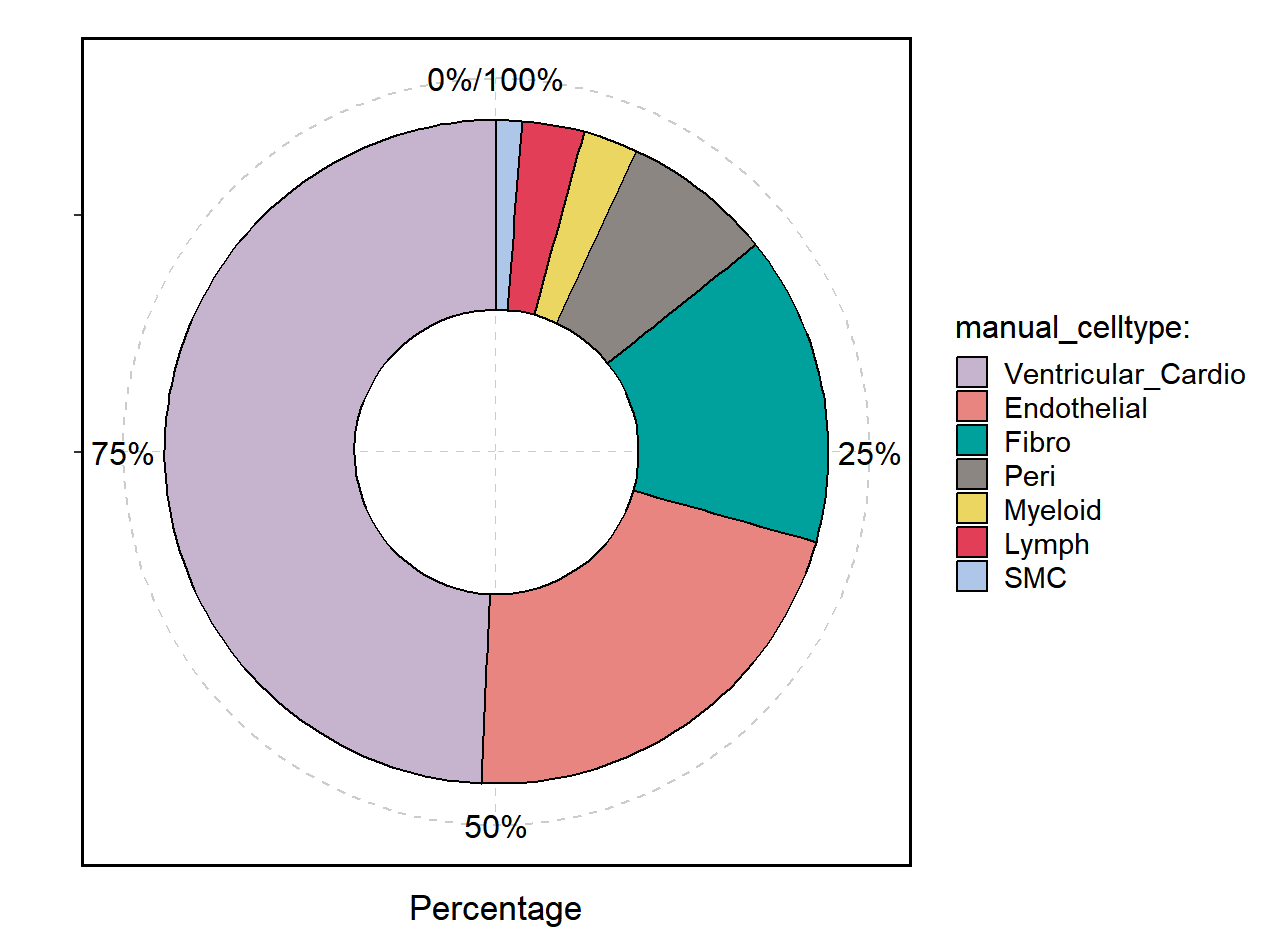

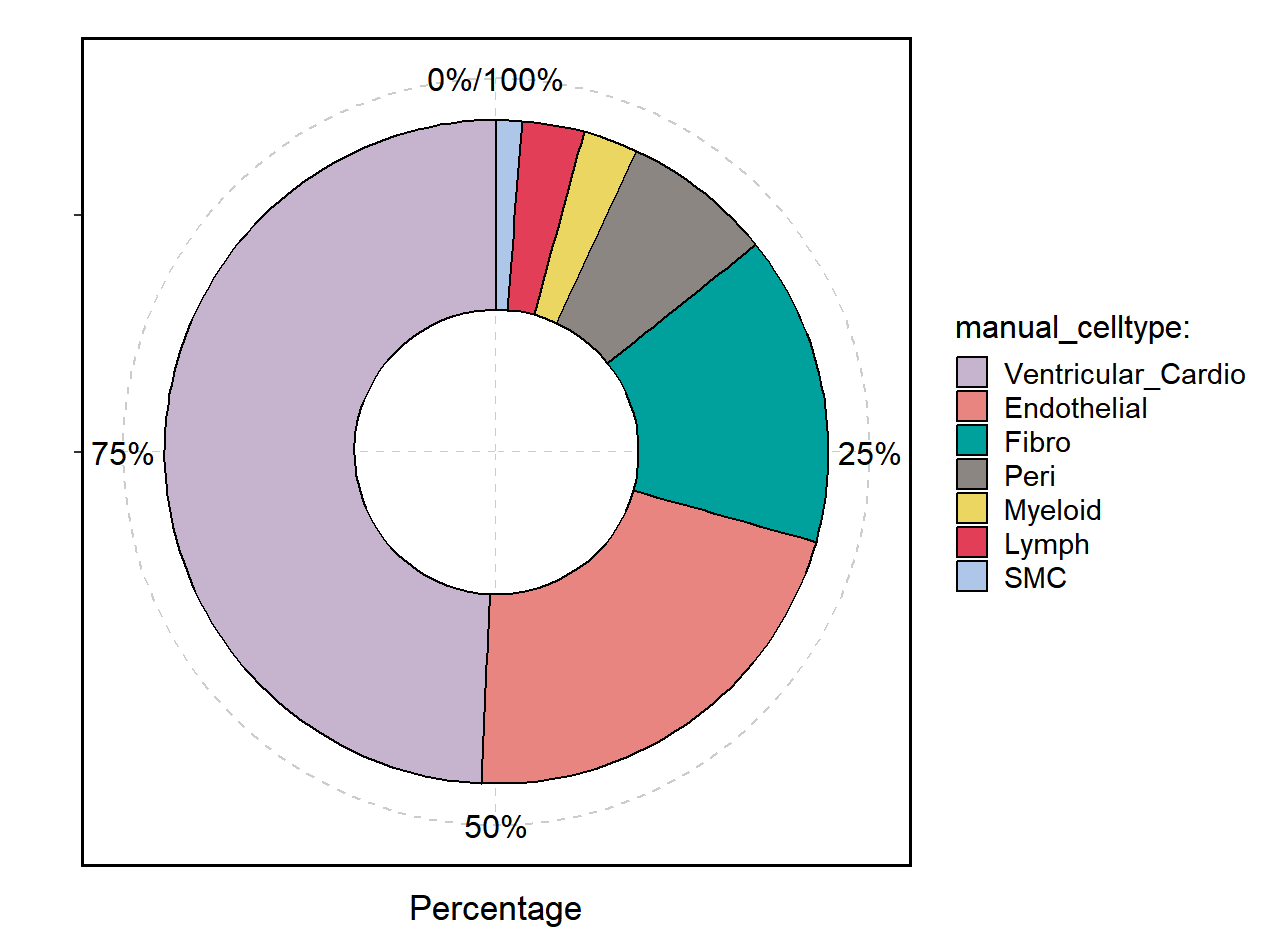

C)
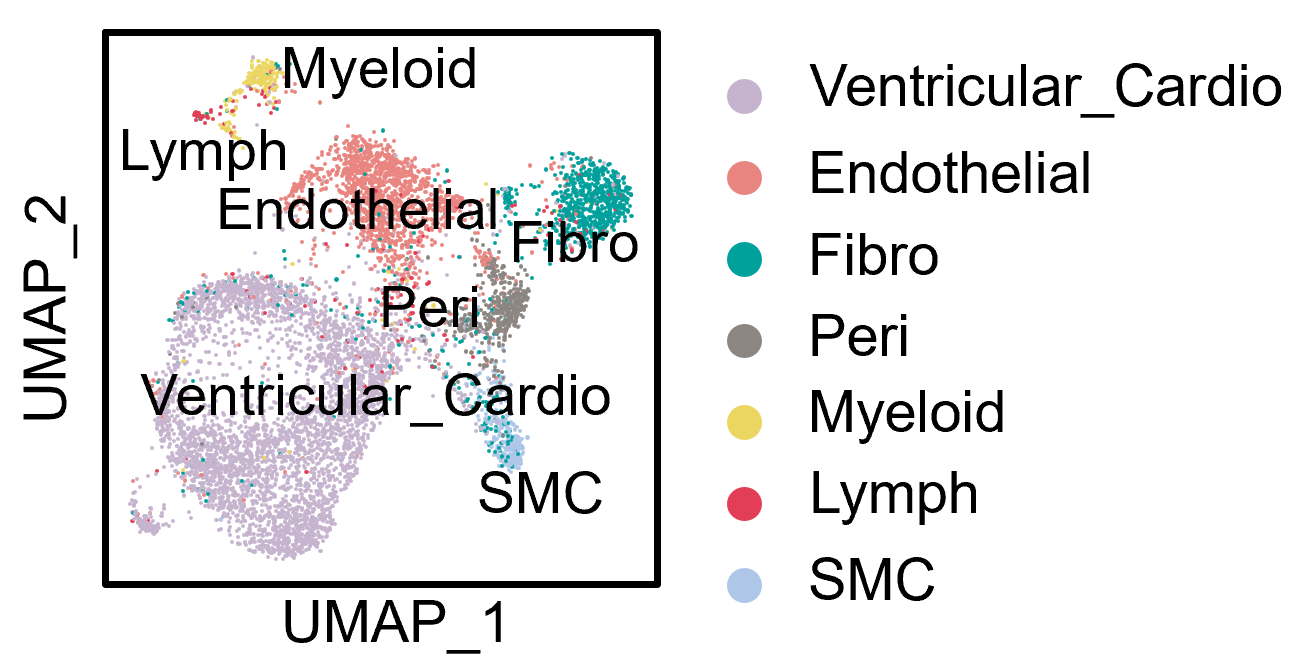
Interventricular Septum UMAP (left) and pie chart of cell types (right)

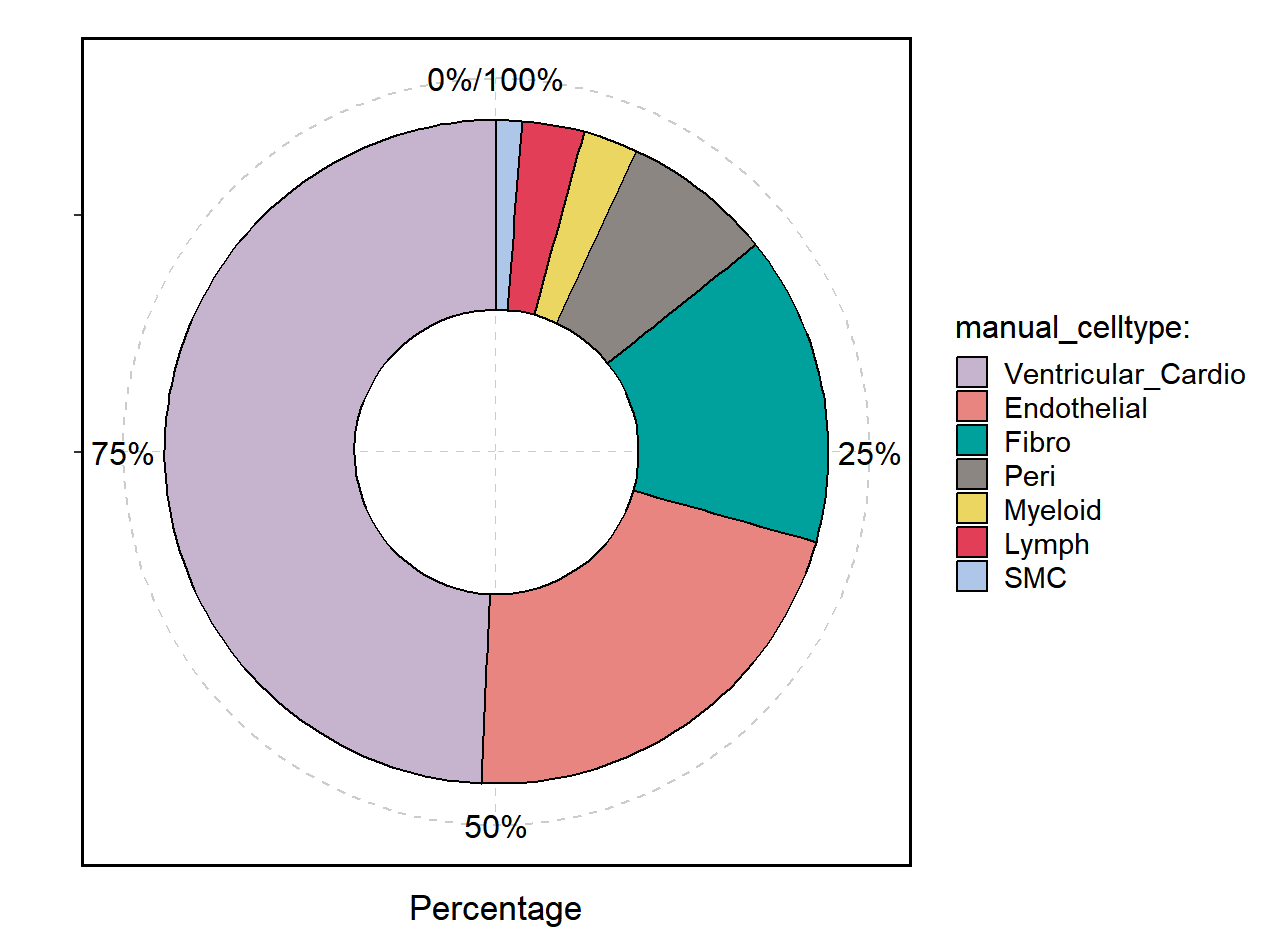

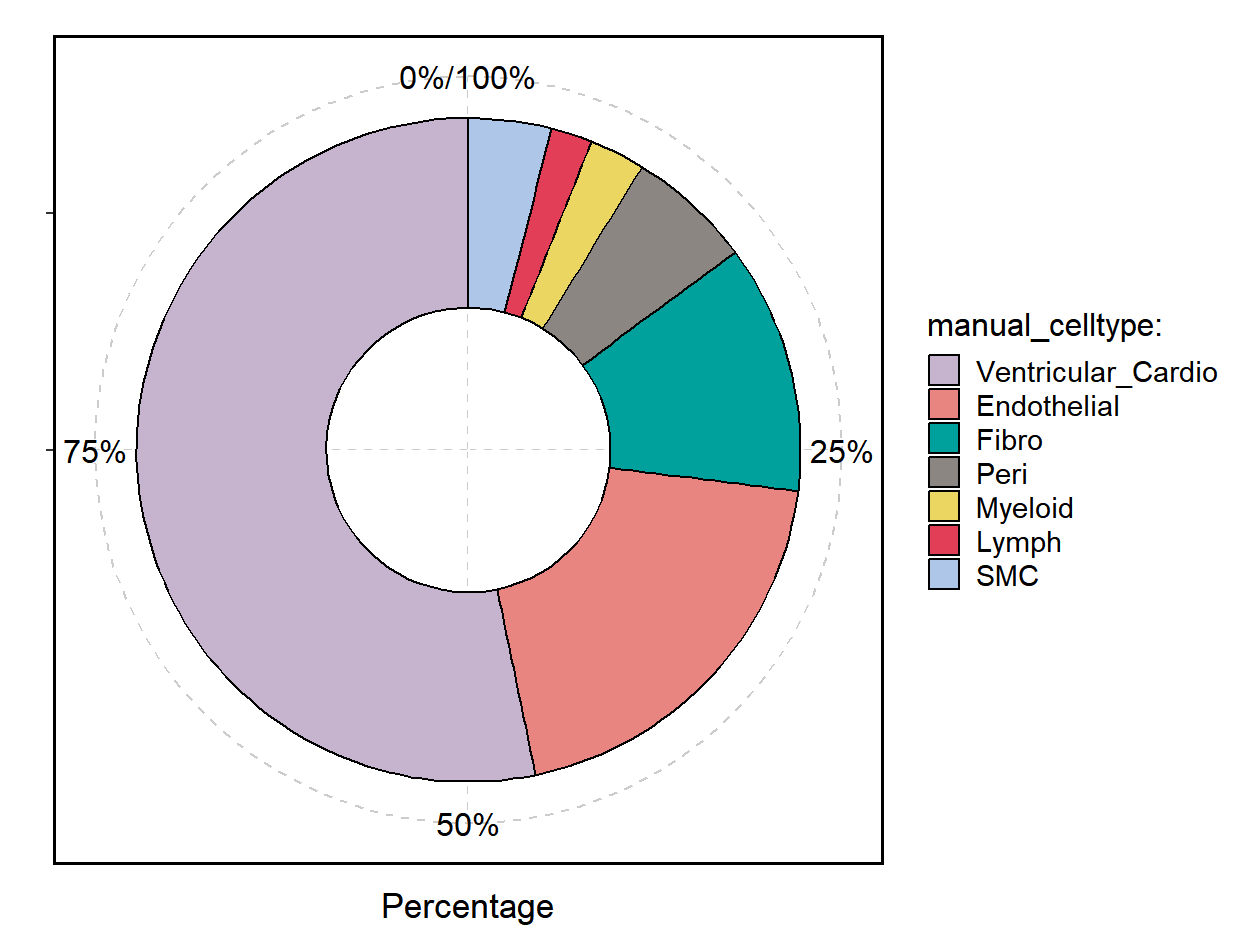

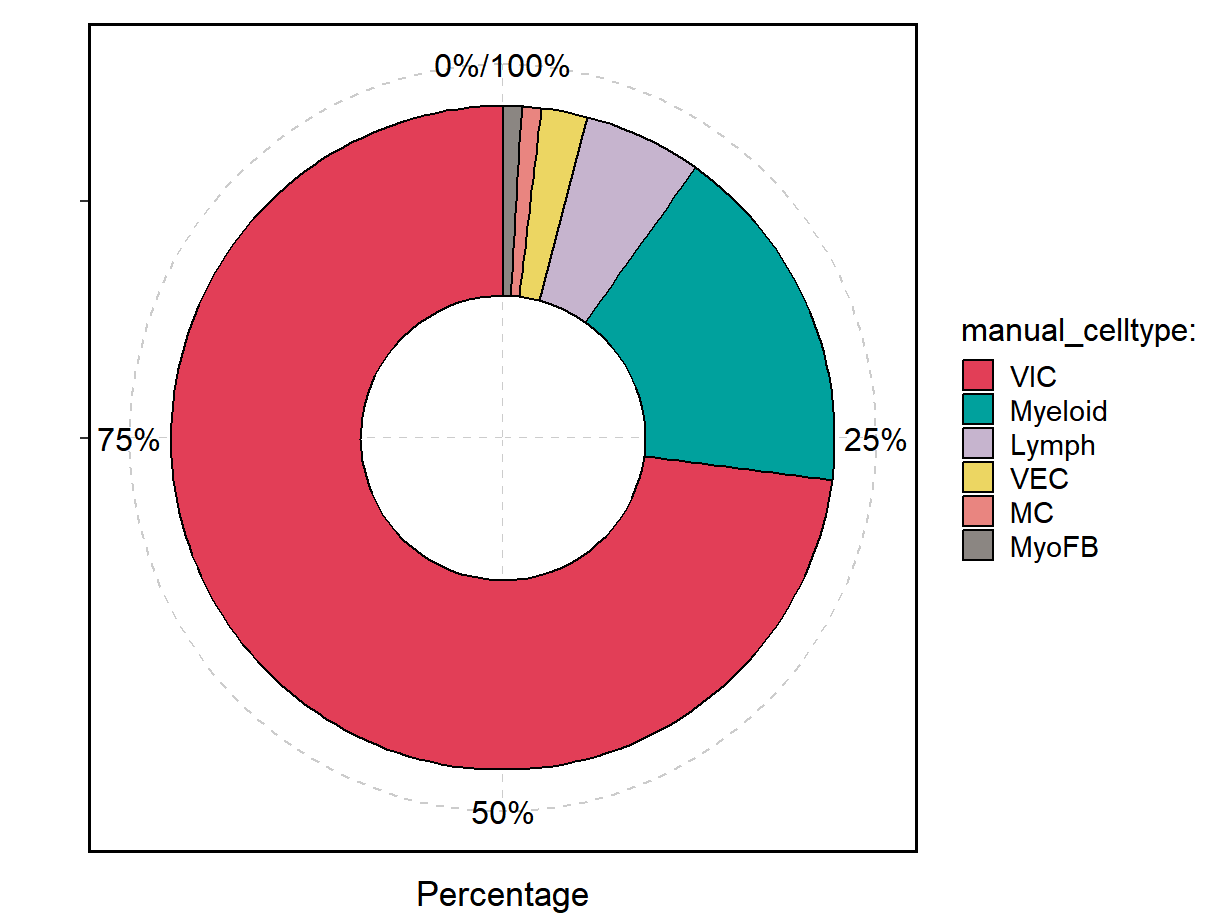

D)
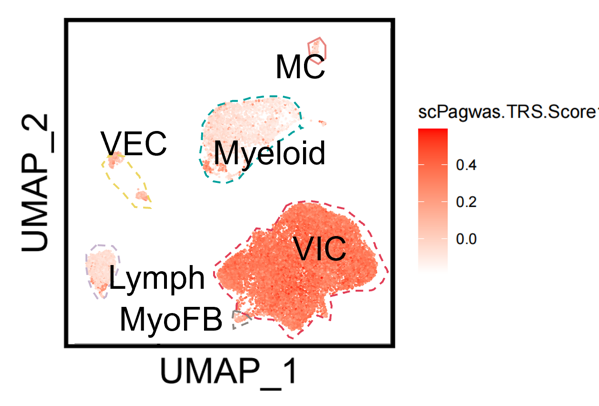
Mitral valve tissue scPagwas
E)
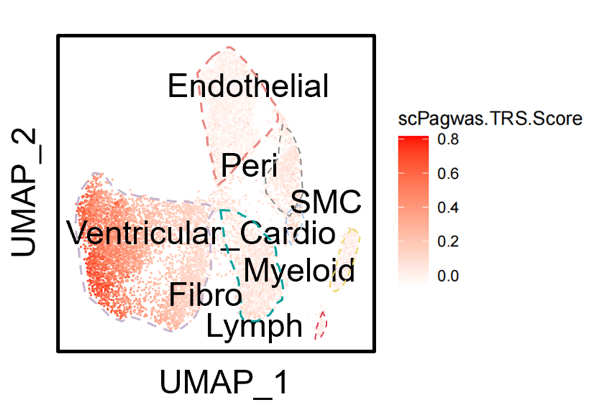
Papillary muscle scPagwas
F)
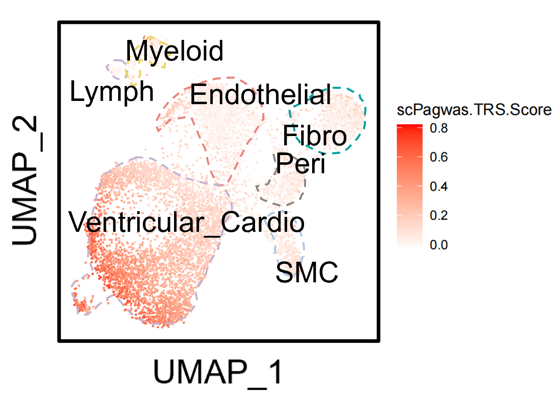
Interventricular septum scPagwas

**Legend:** Uniform manifold approximation and projection plots (left) and pie chart of cell types (right) for (A) mitral valve tissue, (B) papillary muscle, and (C) interventricular septal tissue. scPagwas results for (D) mitral valve tissue, (E) papillary muscle, and (F) interventricular septum. Darker red in scPagwas reflects stronger disease-associated cell type association. Abbreviations as follows: Lymph (lymphocyte); MC (mast cell); Myeloid (myeloid cell); MyoFB (myofibroblast); VEC (valve endothelial cell); VIC (valve interstitial cell); Ventricular_Cardio (ventricular cardiomyocyte); Peri (pericyte); Fibro (fibroblast); SMC (smooth muscle cell).

**Supplemental Figure 8:** Histological evaluation of interventricular septal and papillary tissue

A) Septal tissue

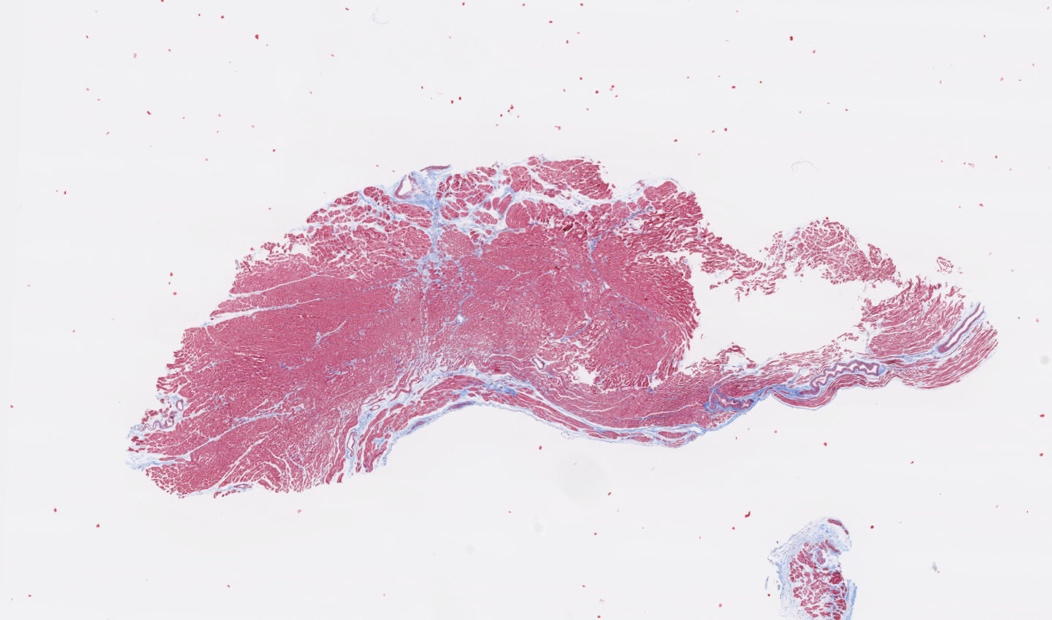

B) Papillary muscle tissue

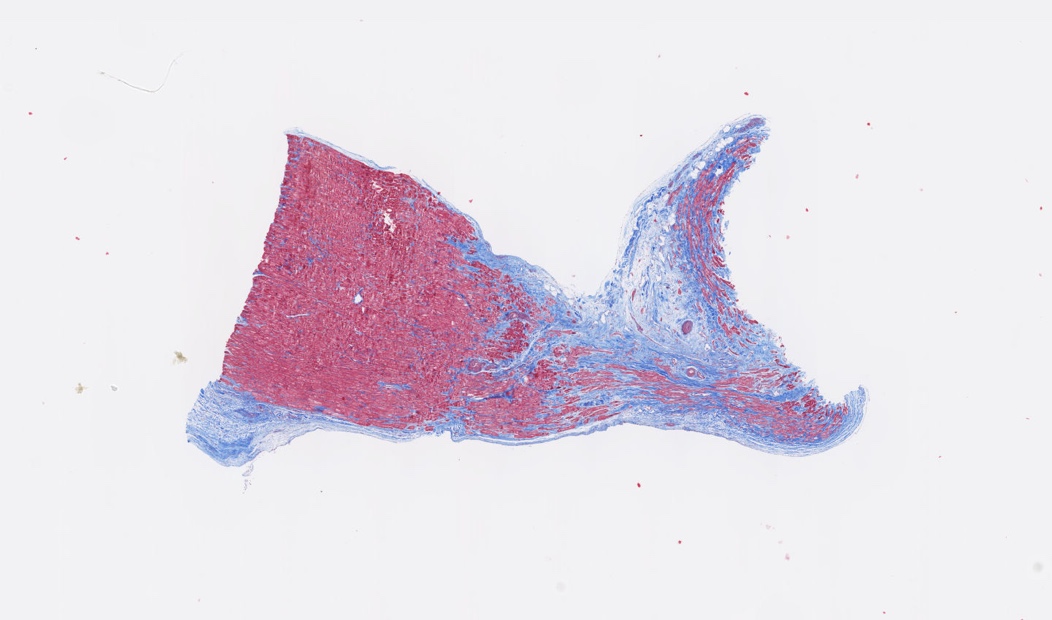

**Legend:** Histological samples with Mason’s trichrome staining of (A) septal cardiovascular tissue and (B) papillary muscle tissue. Blue staining represents collagen/fibrosis.

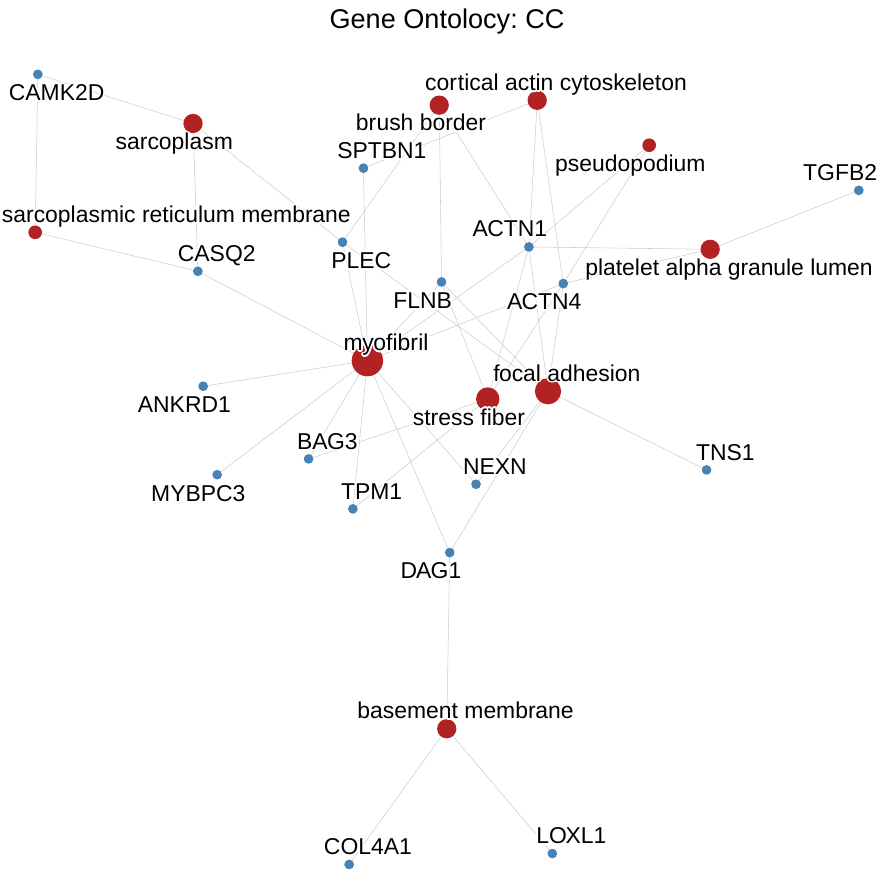
**Supplementary Figure 9:** Gene ontology concept network plot for 50 genes prioritized by mitral valve prolapse GWAS and differentially expressed between fibrosis and no-fibrosis samples

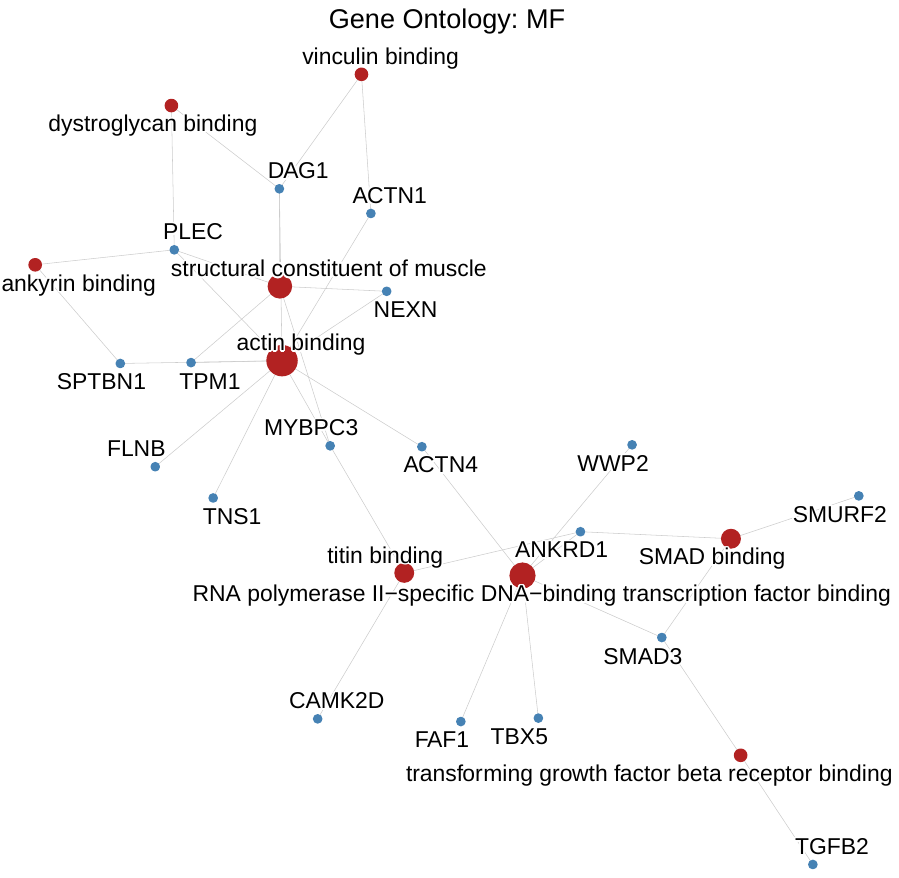

C)
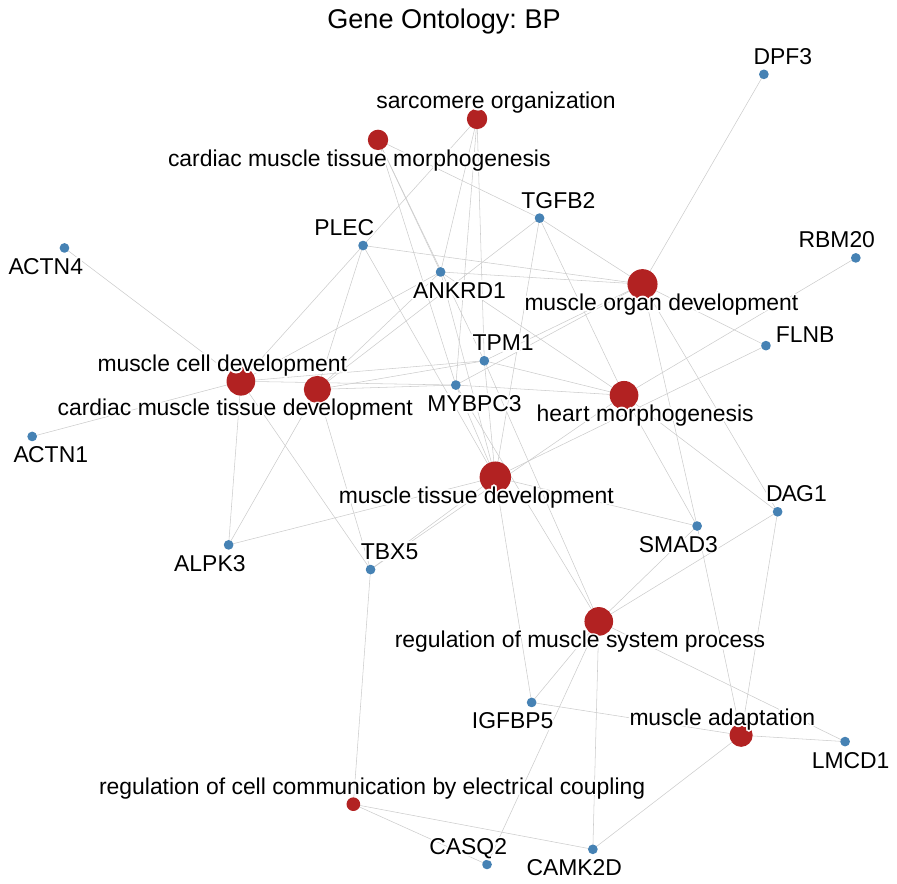

**Legend:** Concept network plot of significant gene ontologies for biological processes (A), cellular components (B), and molecular functions (C) prioritized by mitral valve prolapse GWAS and significantly differentially expressed between papillary muscle (fibrosis) and intraventricular septum (no fibrosis). Size of nodes indicates number of contributing genes.

**Supplemental Figure 10:** P-P plot of MVP GWAS and myocardial fibrosis GWAS

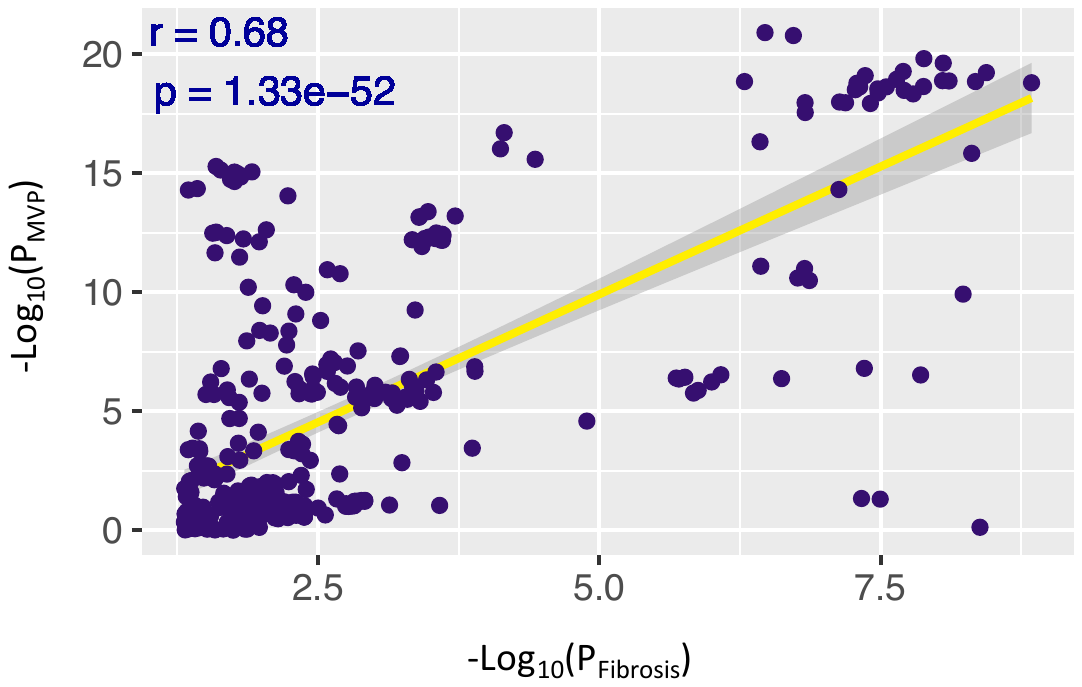

**Legend:** P-P scatterplot of -Log_10_ transformed P-values for our MVP GWAS (Y-axis) and recently published interstitial myocardial fibrosis GWAS^2^ (X-axis). Correlation coefficient (r) and P-value for correlation are displayed at upper left corner of plot. Abbreviations as follows: MVP (mitral valve prolapse); P (P-value); r (Pearson’s correlation coefficient).

**Supplemental Figure 11:** PheWAS of ECG traits for rs17046126 (MVP lead variant prioritized to *CAMK2D*)

rs17046126:

MVP Risk locus prioritized to *CAMK2D*

ECG effect allele: C

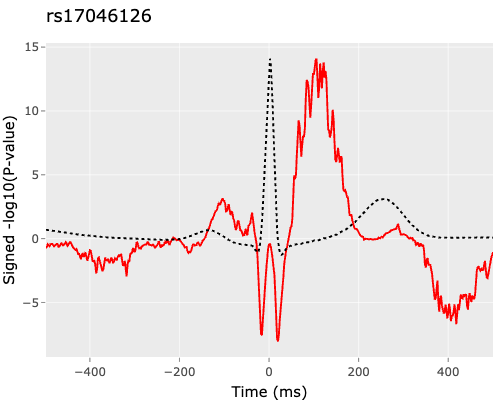

**Legend:** PheWAS of ECG associations (time in milliseconds from the QRS peak) of rs17046126 (intronic to *CAMK2D*). Dotted line represents typical ECG waveform. Red line represents -log10(P-value) for association between variant and time to QRS peak.

**Author Funding and Acknowledgements**:

A.G. received support from Agence Nationale de la Recherche (ANR-23-CE17-0004-01_ACT). An.G. is founder of Real World Genetics Oy. M.Y. is funded by National Natural Science Foundation of China (No. 82400439). J.M. is funded by NIH grant HL167482. R.D. reported being a scientific co-founder, consultant, and equity holder for Pensieve Health (pending) and being a consultant for Variant Bio and Character Bio. R.D. is supported by the National Institute of General Medical Sciences of the NIH (R35-GM124836). M.D. is supported by Forderverein Deutsches Herzzentrum. G.P. is supported by R01HL127564 from the NHLBI. S.K. is supported by K99HL169733 from NHLBI. P.N. reports research grants from Allelica, Amgen, Apple, Boston Scientific, Cleerly, Genentech / Roche, Ionis, Novartis, and Silence Therapeutics, personal fees from AIRNA, Allelica, Apple, AstraZeneca, Bain Capital, Blackstone Life Sciences, Bristol Myers Squibb, Creative Education Concepts, CRISPR Therapeutics, Eli Lilly & Co, Esperion Therapeutics, Foresite Capital, Foresite Labs, Genentech / Roche, GV, HeartFlow, Magnet Biomedicine, Merck, Novartis, Novo Nordisk, TenSixteen Bio, and Tourmaline Bio, equity in Bolt, Candela, Mercury, MyOme, Parameter Health, Preciseli, and TenSixteen Bio, royalties from Recora for intensive cardiac rehabilitation, and spousal employment at Vertex Pharmaceuticals, all unrelated to the present work. P.N. is supported by R01HL127564 and R01HL127564-07S1 from National Heart, Lung, and Blood Institute (NHLBI). R.L. is supported by NIH R01HL173930, American Heart Association grants 22TPA963793 and 24CSA1255237, the Ellison Foundation and Leducq Foundation grant 22ARF02 for the Preventing Rheumatic Injury bioMarker Alliance. Copenhagen Hospital Biobank is supported by the Department of Clinical Immunology, Rigshospitalet, Copenhagen University Hospital, Copenhagen, Denmark and by grants from Novo Nordisk Foundation (NNF23OC0082015, NNF17OC0027594) and Rigshospitalet Research Council (Framework grant). The Danish Blood Donor Study (DBDS) is funded by an annual grant from Bio- and Genome Bank Denmark. The initiation of DBDS was supported by the Danish Administrative Regions (02/2611) and the Danish Council for Independent Research (09–069412). Additionally, the DBDS is funded by the Novo Nordisk Foundation (NNF23OC0082015, NNF17OC0027864, and NNF17OC0027594). J.G. is supported by AUFF Research Grant (47951). C.E. has received unrestricted research grants from Novo Nordisk (administered by Aarhus University) and Abbott Diagnostics (administered by Aarhus University Hospital). He has received no personal fees. M.G.L. receives grant funding from the Doris Duke Foundation (award 2023-0224) and US Department of Veterans Affairs Biomedical Research and Development ( IK2-BX006551), grant funding to the institution from MyOme, and consulting fees from BridgeBio. This research includes data from the Million Veteran Program, Office of Research and Development, Veterans Health Administration, and was supported by MVP000 as well as award #BX004821. This publication does not represent the views of the Department of Veteran Affairs or the United States Government.

**References**

1.	Assmann G. Simple Scoring Scheme for Calculating the Risk of Acute

Coronary Events Based on the 10-Year Follow-Up of the

Prospective Cardiovascular Münster (PROCAM) Study. *Circulation*. 2002;105.

2.	Nauffal V, Di Achille P, Klarqvist MDR, Cunningham JW, Hill MC, Pirruccello JP, Weng LC, Morrill VN, Choi SH, Khurshid S, Friedman SF, Nekoui M, Roselli C, Ng K, Philippakis AA, Batra P, Ellinor PT, Lubitz SA. Genetics of myocardial interstitial fibrosis in the human heart and association with disease. *Nat Genet*. 2023;55:777-786. doi: 10.1038/s41588-023-01371-5
